# EDIT-B Consortium: An External Pan-European Validation of an Innovative Machine Learning-Based Test for Bipolar Disorder Diagnosis

**DOI:** 10.1101/2025.11.05.25339583

**Authors:** Dinah Weissmann, Nicolas Salvetat, Christopher Cayzac, Francisco Checa, Diana Vetter, Francisco Santos-Schneider, Lara Walczer-Baldinazzo, Giovanna Ruggeri, Maurizio Ferrari, Andrea Miranda Mendizabal, Victor Chavarria, Oscar Soto-Angona, Juan Zambrano, Jeff Zarp, Anna Gimenez-Palomo, Marc Valenti, Josep Maria Haro, Lars Kessing, Chantal Henry, Eduard Vieta

## Abstract

**Background:** Differentiating bipolar disorder (BD) from major depressive disorder (MDD) during major depressive episodes remains a significant challenge. EDIT-B is an in vitro diagnostic test based on machine learning (ML) method integrating clinical metadata with adenosine-to-inosine (A-to-I) RNA editing signatures in eight genes (*GAB2*, *IFNAR1*, *IL17RA*, *LYN*, *MDM2*, *PRKCB*, *PTPRC*, *ZNF267*) to differentiate BD from MDD.

**Objective:** The objective of this study is to assess and confirm the diagnostic performance of EDIT-B in a new, independent, double-blind, external multicenter European cohort.

**Methods:** We evaluated EDIT-B results across four European centers (Spain, France, Denmark), compared to physician diagnoses. Explainability and sensitivity analyses were performed to identify the primary drivers of the model.

**Findings:** In 393 patients with current major depressive episode (238 MDD; 155 BD), EDIT-B demonstrated robust performance: AUC-ROC of 0.873 (95%CI: 0.837-0.909); sensitivity of 82.6% (95%CI: 75.7-88.2%); and specificity of 80.3% (95%CI: 74.6-85.1%). Performances were consistent across sites and all stratified patient subgroups, corroborating previous results. Explainability analysis pointed RNA editing biomarkers as the most important variables for EDIT-B diagnosis.

**Conclusion:** This study confirms the diagnostic performances of EDIT-B showing that A-to-I RNA editing coupled with ML methods is a reliable approach to distinguish BD from MDD.

**Clinical implications:** EDIT-B represents an innovative, reliable, and complementary diagnostic tool supporting psychiatric practice and enhancing clinical outcomes with potential impact in reducing the diagnostic delays, tailoring the therapeutic strategy and improving therapeutic alliances. These findings may represent a significant advance toward precision psychiatry, with potential to reduce misdiagnosis and inappropriate treatment.

**KEY MESSAGES:** *What is already known on this topic:* Bipolar disorder (BD) is frequently misdiagnosed as major depressive disorder (MDD) due to overlapping depressive symptoms, leading to diagnostic delays of up to a decade and important clinical consequences. Current diagnostic approaches rely on clinical assessment and lack specificity while few objective/biologically-based tools are available for accurate early differential diagnosis.

*What this study adds:* This study corroborates EDIT-B test performances for diagnosing BD during depressive episodes in a large, independent European multicenter replication cohort. This study also highlights the importance of RNA editing biomarkers for EDIT-B diagnosis. Sensitivity analyses show robust performances and no major difference among subgroups in all tested conditions including comparison between untreated to treated patients for several classes of psychiatric medications.

*How this study might affect research, practice or policy:* EDIT-B addresses the need of a reliable and objective diagnosis of bipolar disorder during depressive episodes. The EDIT-B test represents a significant advance in precision psychiatry with potential to minimize misdiagnosis and diagnostic delays, to accurate and tailor treatment strategies.

## INTRODUCTION

The differential diagnosis of bipolar disorder (BD) and major depressive disorder (MDD) remains a critical unmet need. Although both conditions are characterized by depressive episodes, their pathophysiological and treatment trajectories differ substantially. BD often initially presents with depression, leading to frequent misdiagnosis as MDD and an average diagnostic delay of nearly a decade[1,2]. This delay has important clinical consequences, including inappropriate pharmacotherapy, increased risk of treatment-emergent mania, and suicide[3–5]. Traditional diagnostic approaches rely on symptomatology and structured interviews, lacking the precision to disentangle overlapping phenotypes[6]. While certain clinical features (e.g. early age at onset, mood lability and family history) may suggest BD, their predictive value remains limited. The limited availability of objective, biologically grounded tools for differential diagnosis impedes early and accurate identification of BD.

Recent advances in molecular psychiatry have focused on immune and oxidative pathways[7]. Among emerging candidates, RNA editing, a dynamic mechanism modifying nucleotide sequences post-transcription, has shown promise. Dysregulated editing patterns were reported in BD and other neuropsychiatric disorders, implicating this process in mood regulation and neuroinflammation[8,9].

In a previous study, EDIT-B, a blood diagnostic test, showed high sensitivity and specificity for distinguishing BD from MDD during major depressive episodes (MDE)[9]. EDIT-B uses a locked ML algorithm based on 22 RNA editing biomarkers from 8 genes (*GAB2*, *IFNAR1*, *IL17RA*, *LYN*, *MDM2*, *PRKCB*, *PTPRC*, *ZNF267*) and clinical data for performing the diagnostic. The editing biomarkers were selected through a multi-step strategy combining A-to-I RNA-editome discovery, stringent filtering, biological/functional network analyses, analytical and independent clinical validation[9,10]. The biomarkers present interconnected pathways involved in neuro-inflammation, microglial activation, immune signaling, oxidative stress and mitochondrial dysfunction, all implicated in the pathophysiology of BD (Supplementary Figure1)[2]. EDIT-B test is currently CE-marked under the In Vitro Diagnostic Directive and already commercialized in some European countries.

Here, we present results and confirm the performance of EDIT-B in a novel external large-scale, independent, double-blind Pan-European cohort. The EDIT-B project is funded by the European Union (Grant 220628/230125).

## METHODS

### Study design and setting

The EDIT-B consortium study (ClinicalTrials.gov NCT05603819) is a double-blind multicenter cross-sectional study enrolling adults between June 2022 and September 2024 across four clinical sites: two in Barcelona (Spain), one in Paris (France), and one in Copenhagen (Denmark). Reporting followed the SPIRIT guideline (Supplementary Table1). This study adhered to the STARD-AI guidelines for reporting of AI-centered diagnostic test accuracy studies. Written informed consent was obtained from participants. This study was conducted in accordance with the Declaration of Helsinki (2013). An external central laboratory (SYNLAB Italia - Genetic Unit, Castenedolo, Italy) processed the blood samples and perform EDIT-B test on a dedicated secure web platform. SYNLAB was blinded from the clinicians’ diagnoses and clinicians were blinded from EDIT-B results. The protocol of this study has been published in Miranda et al[11].

### Clinical Assessment

A total of 426 participants were screened for eligibility: 259 with MDD, 165 with BD, and two with missing diagnostic. Eight participants did not meet the inclusion/exclusion criteria. Additionally, 14 participants were excluded for protocol deviations and 14 further participants were excluded for intended use or technical quality criteria. The final analytical set included 393 participants: 238 with MDD and 155 with BD (Supplementary Figure2). Eligibility criteria and sample size calculation are described in supplementary methods section.

### EDIT-B In vitro diagnostic test

The panel of pre-mRNA sequences used by EDIT-B includes *GAB2*; *IFNAR1*; *IL17RA*; *LYN*; *MDM2*; *PRKCB*; *PTPRC*; and *ZNF267*. EDIT-B genes were initially identified through an unbiased, data-driven analysis of the blood RNA editome[10].

Details on the assay protocol and on the algorithm were previously described[9] and are strictly the same used in this study. Briefly, blood samples were collected at the clinical centers using PAXgene blood RNA tubes and stored upright at room temperature (18−25°C) for 2 hours before freezing (-20°C). Patients were allowed to eat and to take their routine medicines before blood sampling. Fasting status and the time elapsed since the last medication administration were not documented. The samples were sent on dry ice every two months to Synlab using time- and temperature-controlled transport. Blood samples were treated in batch of randomly distributed patients. RNA was extracted and qualified before PCR, then the PCR product amplification was purified and quantified before indexing and pooling into a final library. The library was denatured, loaded and sequenced at standard concentrations on an Illumina NextSeq 500/550 sequencer (Figure1A).

**Figure 1.**
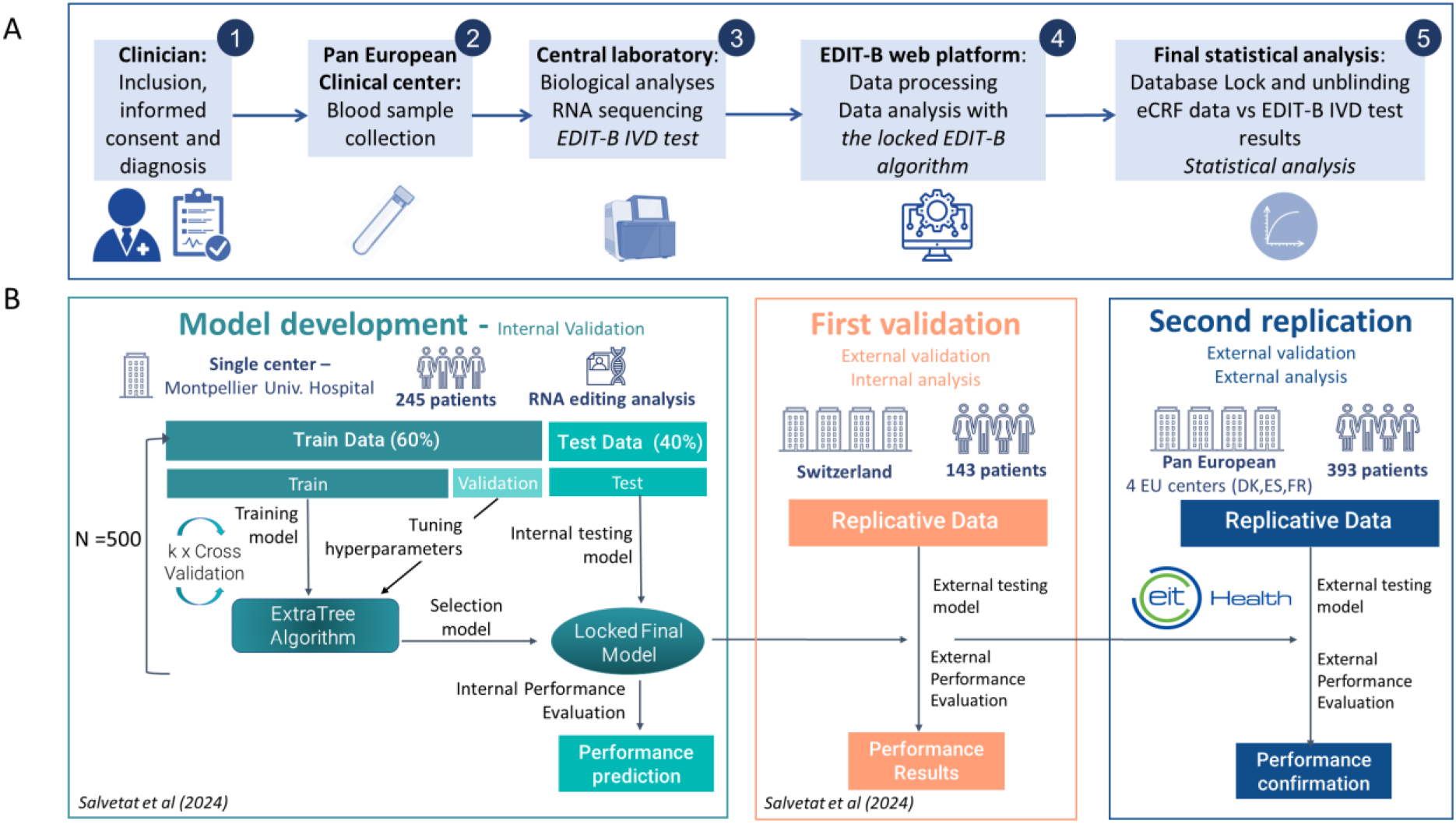
Workflow of the EDIT-B consortium study: From Patient Recruitment to AI Algorithm Validation. A) (1) Participants were enrolled by the clinician and received a diagnostic evaluation; (2) After inclusion, a blood sample was collected at the respective study centre; (3) Samples were shipped to the central laboratory (Synlab-IT) for performs the EDIT-B IVD test; (4) Data were analyzed using the locked EDIT-B algorithm via the HDS EDIT-B web platform (5) Clinical eCRF data and EDIT-B IVD test results were compared in the final statistical analysis of the locked and certified analytical set. B) The EDIT-B algorithm was developed by randomly splitting the model development dataset into 60% training and 40% testing subsets. Preprocessing was applied to the training data, and an ExtraTrees model was built using k-fold cross-validation for hyperparameter optimization. The model was evaluated on the test data, and this process was repeated 500 times with random splits. The final model was selected based on minimizing training error and maximizing AUC-ROC. It was then locked and applied to two external cohorts (Switzerland study n=143 and Pan-European double_blind multicentric study n=393) for validation using the same fixed-function parameters as in internal testing.

Sequencing data were uploaded to the EDIT-B web secure platform, and each patient was processed individually on a Health Data Hosting-certified server using bioinformatic protocol previously detailed[9]. A standardized report with the EDIT-B result was produced for each patient, including sample identifiers, quality control metrics, and algorithm specifications (Figure1A). EDIT-B is a locked ML algorithm with a fixed function based on ExtraTree method using 22 RNA editing biomarkers, demographic data (age, sex), and psychiatric treatments (antidepressants, antiepileptics, antipsychotics (including lithium), anxiolytics, and hypnotics/sedatives, as well as alcohol and tobacco use. Medication was coded according to the WHO Anatomical Therapeutic Chemical (ATC) classification system. The initial development, training and internal validation of the EDIT-B model is shown in Figure1B. The same fixed-function parameters applied to the internal validation were employed in the two external evaluations, ensuring consistency among the cohorts. A description of the AI model development and the associated parameters is detailed in the Supplementary Methods and Supplementary Table2.

### Data analysis

Performance of the test was estimated by sensitivity, specificity and overall accuracy. ROC curves and DeLong’s test (pROC R package[12]) were used to evaluate binary classification performance and compare AUC-ROC values across populations. Precision-Recall (PR) curves and the corresponding Area Under the Curve (AUC-PR) were calculated using the ‘precrec’ R package[13] to evaluate the model’s ability to identify positive instances while minimizing false positives. An optimized cut-off for positive predictive value (PPV), determined in the previous cohort using the cutpointr R package[14] to minimize false positives, was applied to the current study population. To ensure robustness and generalizability, comprehensive stratified sensitivity analyses were performed on the pooled sample from all centers. These evaluations encompassed psychiatric treatment status (Figure3), clinical complexity and comorbidities (Supplementary Figure9–10), demographic and basic clinical characteristics including age and disease onset (Supplementary Figure11-13), disease progression and recurrence markers such as illness duration and depressive episode frequency (Supplementary Figure14–16) and blood collection timing (Supplementary Figure17). Any site-specific harmonization was performed. Variable importance was assessed in EDIT-B model using TreeSHAP[15]. TreeSHAP decomposes predictions into additive feature contributions, quantifying each predictor’s impact on the model output. Global importance was computed as the mean absolute Shapley value across observations, reflecting each variable’s overall contribution. Descriptive statistics summarized demographic and clinical characteristics and effect sizes were calculated using the effectsize R package[16], with two-sided 95% confidence intervals (CIs) reported for all estimates. Model calibration, and Target Editing Index (TEI) methods and other additional analysis are described in supplementary methods.

## RESULTS

### Patient demographic and clinical characteristics

Patients were matched for sex, age, and severity of depression based on Montgomery-Åsberg Depression Rating Scale (MADRS) score, with no significant between-group differences. The proportion of female patients was similar between the MDD and BD groups (61.8% vs. 61.3%, respectively; p-value=1). The great majority of patients (90.8%) were identified as White. No significant differences in ethnic distribution were observed between patients with MDD and those with BD. Body mass index (BMI) was comparable across groups. As expected, depression severity assessed by MADRS score was not statistically significantly different (p-value=0.9) between both groups (mean score of 32.1 for MDD patients and 32.2 for BD patients). In contrast, BD patients had statistically significantly higher Young Mania Rating Scale (YMRS)scores than major depressive disorder patients (p-value<0.001), even their YMRS score stays very low (1.1 for the MDD vs 1.9 for the BD). Regarding psychotropic treatments, approximately 79 % of the patients were treated with antidepressants, 63 % with antipsychotics, 35 % of patients received anxiolytics, while 21 % of patients received hypnotics/sedatives and 35 % received antiepileptics. Smoking was reported by 37% of patients, and alcohol abuse by 8%, with no significant between-group differences (p-value=1) (Table1). Descriptive statistics, including sample sizes and depression severity, were also performed by recruitment sites (Supplementary Table3).

**Table 1:** Demographic and clinical characteristics of patients included in the Analytical Set. Data are the mean ± SEM. p-values of main characteristics are obtained with the student’s t-test or chi-squared test. MDD: Major depressive disorder patients; BD: bipolar disorder patients; MADRS: Montgomery-Åsberg Depression Rating Scale; YMRS: Young Mania Rating Scale; BMI: Body Mass Index. ᵃCohen’s d. Benchmarks: |d| ≈ 0.2 small, ≈ 0.5 medium, ≈ 0.8 large. ᵇOdds ratio. ᶜCramér’s V. Benchmarks: ≈ 0.1, small; ≈ 0.3, medium; ≈ 0.5, large.

|  | Analytical Set Cohort |  |  |  |  |
| --- | --- | --- | --- | --- | --- |
|  | Total | MDD | BD | Effect Size | P-value |
| **Number, n** | 393 | 238 | 155 |  |  |
| **Age** |  |  |  |  |  |
| Age (min-max) | 18-79 | 18-79 | 18-79 |  |  |
| Age (mean ±SD) | 46.2 ±15.3 | 46.9 ±15.0 | 45.1 ±15.8 | 0.12<sup>a</sup> [-0.08, 0.32] | 0.2 |
| **Sex** |  |  |  |  |  |
| Male (n (%)) | 151 (38.4) | 91 (38.2) | 60 (38.7) | 1.02<sup>b</sup> [0.58, 1.81] | 1 |
| Female (n (%)) | 242 (61.6) | 147 (61.8) | 95 (61.3) |  |  |
| **Race** |  |  |  |  |  |
| Asian | 6 (1.5%) | 3 (1.3%) | 3 (1.9%) |  |  |
| Black | 13 (3.3%) | 7 (2.9%) | 6 (3.9%) |  |  |
| White/Caucasian | 357 (90.8%) | 216 (90.8%) | 141 (91.0%) |  |  |
| Other | 17 (4.3%) | 12 (5.0%) | 5 (3.2%) |  |  |
| All |  |  |  | 0.06<sup>c</sup> [0.00, 0.12] | 0.7 |
| **BMI, kg/m2 (mean±SEM)** | 26.2 (±5.5) | 26.2 (±5.6) | 26.1 (±5.3) | 0.02<sup>a</sup> [-0.18, 0.22] | 0.9 |
| **Clinical Characteristics** |  |  |  |  |  |
| MADRS score (mean±SEM) | 32.1 (±6.5) | 32.1 (±6.6) | 32.2 (±6.3) | -0.02<sup>a</sup> [-0.22, 0.19] | 0.9 |
| YMRS score (mean±SEM) | 1.4 (±2.0) | 1.1 (±1.8) | 1.9 (± 2.3) | -0.40<sup>a</sup> [-0.60, -0.19] | <**0.001** |
| **Psychotropic Treatments** |  |  |  |  |  |
| Anxiolytics (n (%)) | 139 (35.4) | 100 (42.0) | 39 (25.2) | 0.47<sup>b</sup> [0.26, 0.85] | <**0.05** |
| Hypnotics/Sedatives (n (%)) | 83 (21.1) | 58 (24.4) | 25 (16.1) | 0.59<sup>b</sup> [0.29, 1.20] | 0.2 |
| Antiepileptics (n (%)) | 136 (34.6) | 41 (17.2) | 95 (61.3) | 7.63<sup>b</sup> [3.95, 14.71] | <**0.001** |
| Antipsychotics (n (%)) | 249 (63.4) | 115 (48.3) | 134 (86.5) | 6.86<sup>b</sup> [3.42, 13.74] | <**0.01** |
| Antidepressants (n (%)) | 309 (78.6) | 232 (97.5) | 77 (49.7) | 0.03<sup>b</sup> [0.01, 0.09] | <**0.001** |
| All treatments Classes |  |  |  | 0.35<sup>c</sup> [0.25, 0.43] | 0.2 |
| **Substances Addiction** |  |  |  |  |  |
| Tobacco (n (%)) | 147 (37.4) | 88 (37.0) | 59 (38.1) |  |  |
| Alcohol (n (%)) | 30 (7.6) | 19 (8.0) | 11 (7.1) |  |  |
| All |  |  |  | 0.03<sup>c</sup> [0.00, 0.22] | 1 |

Regarding sociodemographic characteristics, participants were commonly single (42.2%), followed by married (32.1%) and separated/divorced (22.9%). Most lived alone (36.6%), with a spouse/partner (35.6%), or with children (26.2%). Over half reported a disability, predominantly temporary (37.7%) or permanent (14.0%). Educational attainment was high: 91.1% had completed secondary school and 49.6% held a university degree. No significant differences were observed between MDD and BD groups (Supplementary Table4).

The mean duration of illness was comparable between the two groups (around10 years). The onset of disease and the first depressive episode tended to occur earlier in patients with BD (p-value<0.001). Among the BD patients, the first depressive episode typically preceded the first manic/hypomanic episode. BD patients experienced a higher number of depressive episodes compared to MDD patients. Depressive polarity at onset was predominant in BD patients (88.4%). Psychotic symptoms were present in approximately one-third of BD patients, compared with fewer than 10% of MDD patients. A lifetime history among first or second generations of affective disorders was reported by 72.9% of BD patients and 49.2% of MDD patients, while a family history of psychiatric illness was more common in BD patients (81.3%) than in MDD patients (60.5%). A family history among first or second generations of completed suicide was reported by approximately 20% of patients in both groups (Supplementary Table5).

According to the Mini-International Neuropsychiatric Interview (MINI), all participants fulfilled criteria for an ongoing MDE of at least two weeks’ duration, with most reporting a recurrent history. Manic or hypomanic episodes were absent in the MDD group, whereas 40.0% and 63.9% of BD patients reported lifetime manic and hypomanic episodes, respectively. Recent suicide attempts were common in both groups (20–30%). Psychiatric comorbidities included panic disorder (19.1% overall; 26.5% in BD vs. 14.3% in MDD), generalized anxiety disorder (15.0%, similar across groups), psychotic disorders (3.8%, predominantly in BD I), alcohol use disorder (7.6%), substance use disorders (4.3%), obsessive-compulsive disorder (8.4%), and eating disorders (2.0%) (Supplementary Table6).

### EDIT-B test performances

The EDIT-B test was evaluated in this study (n=393) (Figure2A). The empirical ROC curve presents an AUC-ROC of 0.873 (CI 95%: [0.837 - 0.909]) and an AUC-PR of 0.843 (CI 95% [0.798-0.884]; Supplementary Figure3A) for EDIT-B test. The 95% confidence intervals (CIs) for the AUC-ROC of EDIT-B test overlapped with those previously reported for the Swiss external validation (AUC-ROC of 0.904, 95%CI [0.853-0.955]) (Figure2B), confirming statistical consistency (DeLong’s test, p=0.33).

**Figure 2.**
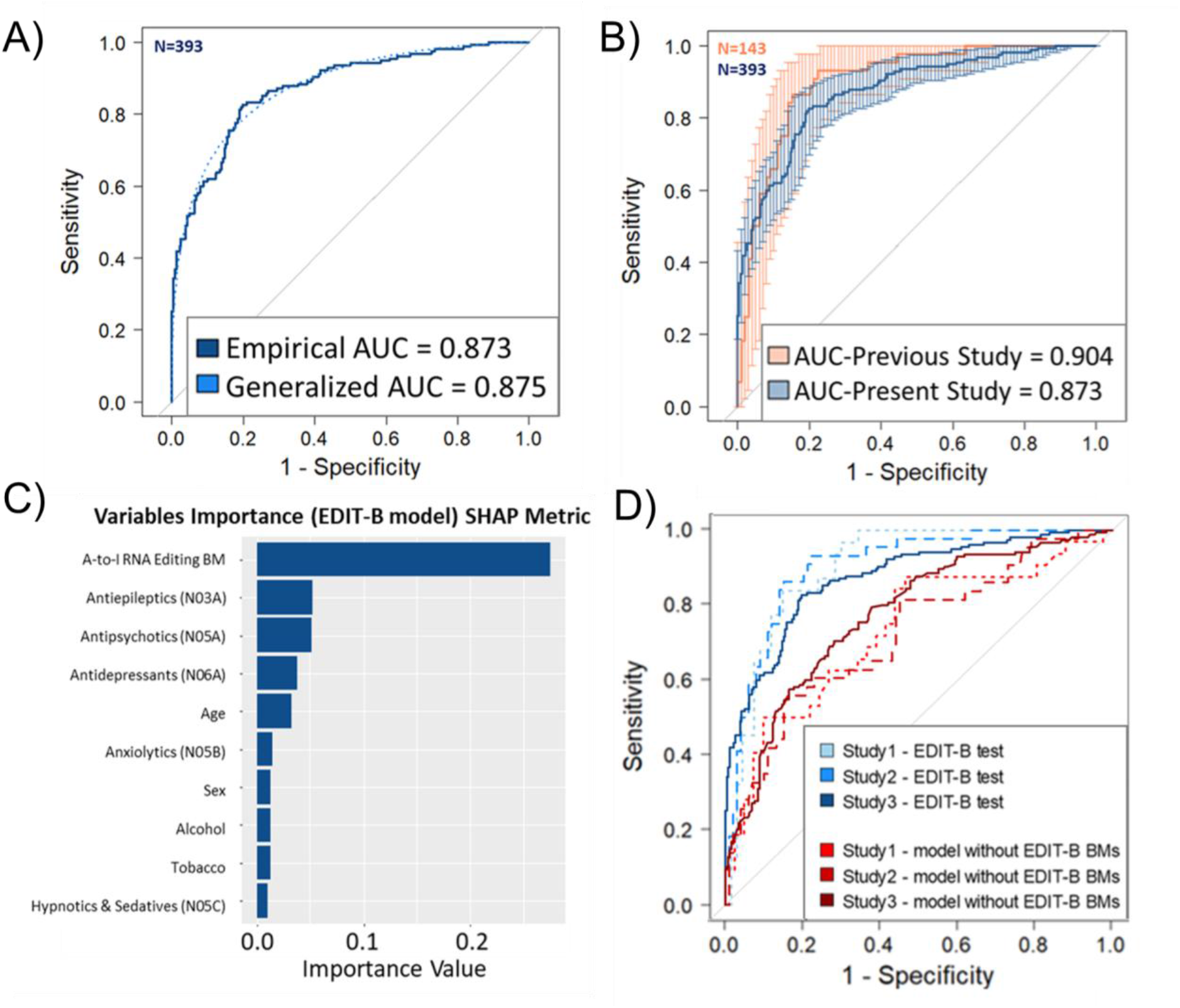
Diagnostic performances, explicability and impact of RNA editing biomarkers of EDIT-B test. A) ROC curves of EDIT-B test in present study (Analytical set, N=393): Light blue line: Generalized Area Under ROC Curve; Dark blue line: Empirical Area Under ROC Curve. B) Comparison of the ROC curves and confidence intervals of EDIT-B test in present study vs previous results [9]: Corail line : previous study ROC curve (Salvetat et al, 2024, N=143); Dark blue line: present study ROC curve (Analytical set, N=393); P-value = p-value of the DeLong’s Test. C) Variable importance using SHAP metric: Bar plot illustrates the importance of variables of mean absolute Shapley values in the EDIT-B test. The length of each bar indicates the relative contribution of each variable to the model’s predictions. The variable “A-to-I RNA Editing BM” exhibits the highest importance, followed by “Antiepileptics (N03A),” “Antipsychotics (N05A),” and other factors. This analysis helps identify key drivers of the model’s performance. D) EDIT-B model development and comparison to clinical variables model excluding EDIT-B biomarkers. ROC curves were generated to illustrate and compare the diagnostic performance of two models, one including RNA editing biomarkers (EDIT-B test) and one excluding them, across the same three cohorts (Development cohort: Study1 (N=98); Switzerland cohort: Study2 (N=143); Current Pan European multicentric cohort - Study3 (N=393)). The inclusion of EDIT-B RNA editing biomarkers statistically improves model performance (DeLong p<0.05), as reflected by higher AUC ROC values in the blue curves (respectively AUC study1=0.901, AUC study2=0.904, AUC study3=0.873) compared to the red curves (respectively AUC study1=0.732, AUC study2=0.723, AUC study3=0.772).

False-positive (MDD patients classified as BD by EDIT-B) and false-negative (BD patients classified as MDD by EDIT-B) cases were also characterized (Supplementary Table7). No significant differences were observed in age of onset, illness duration, psychotic symptoms or frequency of depressive episodes (all p>0.2). Except for alcohol use disorder and the number of mania episodes (p<0.01), psychiatric comorbidities showed no significant association with misclassification (p>0.2).

The diagnostic performances for EDIT-B test (Supplementary Table8) achieved a sensitivity of 82.6% (CI 95%: 75.7%-88.2%), a specificity of 80.3% (CI 95%: 74.6%-85.1%), a negative predictive value (NPV) of 87.6% (CI 95%: 82.5%-91.7%), a positive predictive value (PPV) of 73.1% (CI 95%: 65.9%-79.6%) and an overall accuracy of 81.2% (CI 95%: 77.0%-84.9%). These results confirm the potential use of the EDIT-B test for providing a robust separation between patients with MDD and BD.

Among the four recruiting centers involved in this study, three were BD reference centers (Center 001, Spain; Center 002, Denmark; and Center 003, France). BD reference centers are recognized for their expertise in the management of bipolar disorder, but also treat a broad psychiatric population, including patients with MDD. A detailed center-based analysis was conducted to assess the AUC-ROC and AUC-PR curves of EDIT-B test across the three BD reference centers (Supplementary Figure4A-C and Supplementary Figure5A-C). Overall, EDIT-B test showed high performance across all three BD reference centers (AUC-ROCs of 0.898 (Center 001), 0.887 (Center 002), and 0.893 (Center 003) respectively; AUC-PRs of 0.868 (Center 001), 0.900 (Center 002), and 0.893 (Center 003) respectively). Notably, AUC ROC curves measured in the BD reference centers were superior to the non-BD reference center (Center 004, Spain; AUC of 0.844). When combining data from the three BD reference centers (N=295), EDIT-B test achieved an AUC-ROC of 0.891 and an AUC-PR of 0.892 (CI 95% [0.855-0.927] (Supplementary Figure6A, Supplementary Figure3B and Supplementary Table8) without significant differences from previously[9] reported performance (DeLong’s test, p=0.7, Supplementary Figure6B).

### EDIT-B model assessment

#### A-to-I RNA editing biomarkers

The panel of RNA-editing-based blood biomarkers detailed above was sequenced in the current dataset (n=393). Differential analysis of selected RNA editing biomarkers were summarized in a “Target Editing Index” (TEI) by gene, using the same method and preprocessing than the one applied to the development cohort (initial cohort used to develop and calibrate the EDIT-B algorithm prior to independent validation), showing highly significant discrimination (adjusted p-value<0.05) between MDD and BD populations, demonstrating robust biomarkers performance across cohorts (Supplementary Figure7A-D).

#### Calibration and predictive density

We further assessed the reliability of the probabilistic predictions of EDIT-B using the analytical dataset (n=393). EDIT-B demonstrated robust calibration and predictive performance across the cohort (Brier score=0.19, expected calibration error (ECE)=0.19). The observed agreement between the predicted probabilities and the actual observed outcomes is illustrated in the density and calibration plots (Supplementary Figure8A-B).

#### Feature importance and sensitivity analysis

A global sensitivity analysis was performed to quantify the relative contribution of the variables to diagnostic predictions. SHAP analysis identified biomarker-related variable as the primary drivers of model predictions compared to treatment variables and other clinical factors (age, sex, and addictions; Figure2C).

To assess impact of RNA editing biomarkers on the diagnosis performance, EDIT-B model was compared to a similar model not using RNA editing biomarkers. Briefly, in this analysis we compared: (a) a model without the RNA editing biomarkers including only clinical factors such as age, sex, addiction and psychiatric treatment variables, and (b) EDIT-B, which integrates RNA editing biomarkers with clinical factors and psychiatric treatment features. Both models followed the same pre-processing and machine learning framework across the same three cohorts (Development cohort: Study 1; First Swiss external validation cohort: Study 2; Current second European external validation cohort: Study 3), with discrimination assessed via AUC-ROC as primary performance metric. EDIT-B model achieved the highest AUC-ROC and significantly outperformed the model with age, sex, addictions and treatment alone in the three different cohorts (DeLong p <0.05 for three comparisons) (Figure2D).

Further, to assess a putative confounder effect of medication on EDIT-B performances, a detailed stratified analysis according to psychiatric treatments classes (based on WHO ATC) and patient status (treated vs. untreated) is provided in Figure3; briefly, EDIT-B algorithm performance exhibits no significant differences in the different cases (antidepressants, antipsychotics, anxiolytics and hypnotics & sedatives treatments) except for antiepileptics treatment, for which diagnostic performances (AUC) were higher in untreated patients. The performance of EDIT-B was also evaluated across age, sex and BD subtypes and a detailed subgroup analysis is provided in the Supplementary Figure11-12 and Supplementary Table9. Summarily, algorithm performance was consistent across disease characteristics, psychiatric comorbidities, sex and age groups, while a higher AUC was observed in BD type I as compared with BD type II, although both subtypes maintained high diagnostic accuracy (>80%).

**Figure 3.**
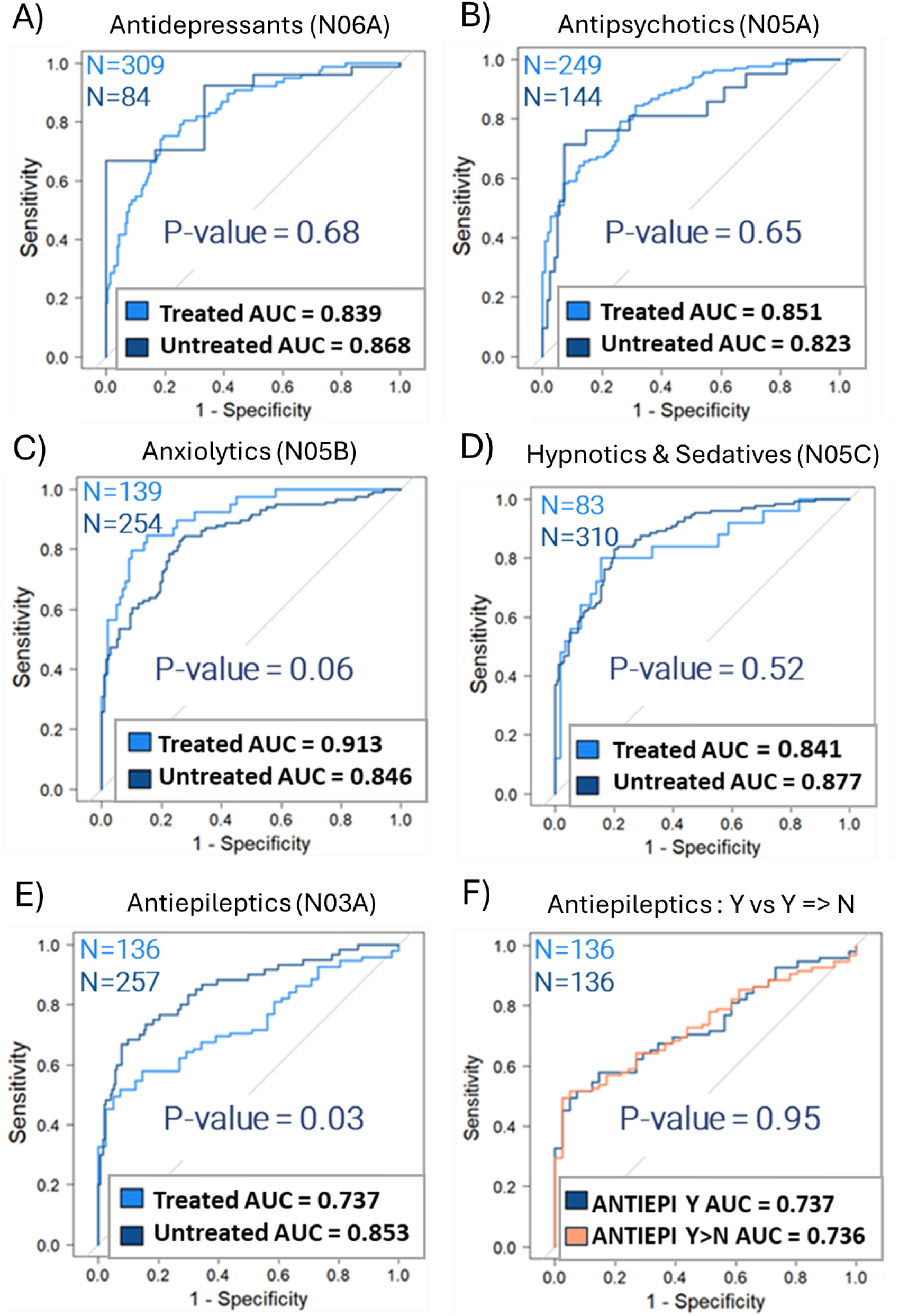
Stratified sensitivity analysis according to psychiatric treatments status. To evaluate the potential medication-related confounding effect, we conducted a stratified sensitivity analysis according to psychiatric treatments status (treated vs. untreated) for (A) Antidepressants treatment (N06A; treated=309 and untreated=84 patients); (B) Antipsychotics treatment (N05A; treated=249 and untreated=144 patients) and (C) Antiepileptics treatment (N03A; treated=136 and untreated=257 patients), (D) Anxiolytics treatment (N05B; treated=139 and untreated=254 patients), (E) Hypnotics & Sedatives treatment (N05C; treated=83 and untreated=310 patients). ROC curves and corresponding AUC values for treated and untreated patients across different psychiatric medication classes are displayed for this analysis, along with the P-values indicating the statistical significance of the differences between the two groups. Notably, the comparison of AUC values for Antiepileptics (DeLong’s p=0.03) is the only one showing a statistically significant difference. (F) The comparison between real antiepileptics uptake (antiepileptic-treated patients) versus simulated no antiepileptic uptake (model is artificially instructed that patients are not taking antiepileptics) demonstrating no significant difference (p=0.95), supporting the robustness of the findings and indicating consistent diagnostic accuracy irrespective treatment exposure. Psychiatric treatments were classified according to the World Health Organization (WHO) Anatomical Therapeutic Chemical (ATC) classification system.

Additionally, diagnostic accuracy remained consistently high in younger populations (aged 18– 25, 18–30, 18-35; AUCs: 0.902, 0.917, 0.877 respectively; all p>0.35) (Supplementary Figure12) and remained consistent regardless of the age of disease onset (≤20 or ≤26 years; p=0.90 or 0.76 respectively) (Supplementary Figure13). Similarly, discriminating power remained stable across illness duration (including early stages ≤1 year; AUC=0.893) (Supplementary Figure14) and regardless of the number of depressive episodes (≤3 or ≤4 episodes; p=0.71 or 0.35) (Supplementary Figure15). Finally, high sensitivity was maintained independently of the frequencies of manic, hypomanic, and mixed episodes, with false-negative rates decreasing as episode frequency increased (Supplementary Figure16). Blood collection timing was homogeneous along the day for both groups, with no significant impact on the performances of the EDIT-B test (p=0.33) (Supplementary Figure17). Together, these findings corroborate the robustness of EDIT-B across diverse clinical trajectories and disease characteristics.

Pearson correlation analyses were performed between the RNA editing levels of biomarkers and various clinical, demographic, and treatment-related covariates. Moderate association (0.2-0.4) were observed between both the number of episodes (manic, hypomanic and mixed) and the age of onset (mania) and RNA editing biomarkers from the genes GAB2, LYN, PTPRC, MDM2, PRKCB, and ZNF267 biomarkers (Supplementary Figure18).

### Diagnostic utility

To evaluate the clinical utility of the model, a decision curve analysis was performed using the analytical dataset. In brief, the model demonstrated a higher net benefit compared to the default strategies of classifying all patients as bipolar or classifying none as bipolar, across a wide range of threshold probabilities (Supplementary Figure19).

## DISCUSSION

This study demonstrates that the innovative EDIT-B test has the potential to be a complementary tool to clinical evaluation, and potential to aid improving the accuracy of the differential diagnosis between MDD and BD. If implemented in clinical routine, a such tool could reduce the diagnostic delay for BD, enabling an earlier patient management and appropriately tailored treatment, reducing the risk of antidepressant-induced mania[17] and ultimately lowering hospitalization rates and associated healthcare costs. Taken together, these results position EDIT-B test as a potentially clinically actionable innovation with direct translational value for personalized and precision psychiatry.

Previous publication demonstrated that using EDIT-B allows to differentiate MDD from BD in two independent cohorts recruited in France and Switzerland. In the present study, we extend this work by evaluating the EDIT-B test (consisting in a locked model, using exactly the same panel and algorithm/parameters) in a third, large and independent European multicenter cohort spanning three countries (France, Spain and Denmark). The current study validated EDIT-B test performances, combining high discriminative performance (accuracy, sensitivity and specificity>80%) with strong reproducibility across diverse clinical settings. When performed on the 393 patients from four centers in three countries, EDIT-B closely replicated the previous results from the original validation study[9], underscoring its robustness across independent cohorts. Moreover, the observed performances were comparable in the great majority of stratification analyses, including psychiatric treatment, age of onset, illness duration, number of depressive/manic episodes, comorbidities, disease characteristics, BD subtype, and sex. Remarkably, age-stratified analyses demonstrate diagnostic performance consistent across all age groups. This finding suggests the potential of the test for early clinical implementation. Altogether, these data support both EDIT-B clinical reliability and its potential for broad generalizability in real-world practice.

When analysis was restricted to patients clinically diagnosed at BD reference centers, EDIT-B performance was greater than that observed for the total population. There is a slight difference in concordance between the specialized reference centers for BD and the general psychiatry center. Indeed, a lower Positive Predictive Value (PPV) indicates a higher proportion of false positives in this center, which slightly reduces the overall accuracy of the model. It is crucial that the diagnosis established by clinicians be as reliable as possible, since this test compares the clinical diagnosis with the EDIT-B test. This finding highlights the usefulness of such a tool as a diagnostic aid, particularly in general psychiatry centers.

Regarding the study population, participants with MDD and BD exhibited largely comparable baseline clinical characteristics, including age, sex, race, BMI, psychiatric treatment, suicidality, suicide behavior, and comorbidities, with the exception of a higher prevalence of panic disorders in BD. This consistency confirms that the recruitment process rigorously adhered to predefined inclusion criteria, ensuring balanced group comparisons. Moreover, while the mean duration of illness was comparable across groups, BD tended to manifest earlier, with the first depressive episode typically preceding the onset of mania or hypomania. In this line, BD patients experienced a higher lifetime number of depressive episodes as compared to MDD patients[18]. Family history patterns also revealed notable differences. A psychiatric family history was more prevalent in BD than in MDD, confirming the strong heritable component of bipolar disorder, reinforcing its genetic underpinnings. Taken together, these findings underscore the complexity of mood disorders and the challenges of differentiating BD from MDD based solely on clinical features during MDE.

Based on clinical features, distinguishing a MDE as part of BD versus MDD remains relatively subjective. This contributes to delays in accurate diagnosis, with the clinician’s experience and expertise playing a crucial role in improving diagnostic accuracy. In primary care, the Mood Disorder Questionnaire has demonstrated a sensitivity around 70% in psychiatric outpatients[19], and good accuracy for screening BD. In the same way, some studies indicate that for patients with bipolar spectrum illness, the sensitivity reaches 70% only for BD type I[20]. Meanwhile, scales such as the 32-item Hypomania symptom check-list, first revision, have shown utility in detecting hypomanic symptoms[21], which can further aid in identifying at risk patients. This confirms the need for additional tool for the differential diagnosis of BD and MDD.

The biological plausibility of the EDIT-B signature is supported by the convergence of its constituent genes on pathways involved in immune signaling, neuroinflammation, oxidative stress and mitochondrial dysfunction. Notably, *IFNAR1*, *IL17RA*, *PTPRC* (also known as *CD45*), *LYN*, *GAB2* and *PRKCB* participate in interconnected signaling networks regulating microglial activation and inflammatory responses through the PI3K-AKT, NF-κB and MAPK pathways[22–26], while *MDM2* is associated with oxidative stress and mitochondria homeostasis via the AKT-p53 axis[27,28]. The relationship between *ZNF267* and the proposed neuroimmune signaling axis remains more elusive. As a transcription factor, *ZNF267* may modulate the expression of genes related to mood disorder and neuroinflammatory responses[29]. Together, these findings support the hypothesis that the EDIT-B signature reflects coordinated neuroimmune and oxidative stress mechanisms implicated in mood disorders.

EDIT-B puts on evidence the potential use of molecular signatures to elucidate the biological underpinnings of depression and bipolar disorder. In this sense, feature importance and sensitivity analyses presented here highlight the key contribution of RNA editing biomarkers to BD diagnosis. The integration of RNA sequencing and ML is emerging as a powerful approach to enhance diagnostic precision in medicine, enabling the identification of robust disease-specific molecular signatures with high precision. This approach underscores the transformative potential of AI-driven genomics and epigenomics in improving clinical outcomes, including for complex disorders in psychiatry, where epitranscriptomic signatures such as the EDIT-B test show great promise.

### Limitations

The present study has some limitations. First, the majority of participants were White/Caucasian, reflecting the demographic profile of the recruitment centers. It may limit the generalizability of our findings to more diverse populations. Second, the patients enrolled in this study were moderately or severely depressed limiting the interpretation of EDIT-B results for the low severity depression states. Regarding the different bipolar sub-classifications (i.e. BD-I vs. BD-II), diagnostic performance varied between BD-I and BD-II in distinguishing MDD from BD. Nonetheless, both subtypes achieved high diagnostic accuracy (>80%). Third, while EDIT-B results demonstrated no significant differences in most stratified sensitivity analyses across medication subgroups, a significant difference was observed in the analysis of patients treated with antiepileptic drugs. However, the comparison between real antiepileptic uptake in treated patients and simulated no antiepileptic uptake (in which the model was artificially instructed to consider the patients were not taking antiepileptics) showed no significant difference (p=0.95). Fourth, the relatively high cost of the NGS-based test may restrict its availability in clinical settings. Fifth, due to the real-world design of the study, detailed medication timing and fasting status were not recorded; however, subgroup analysis by sampling time suggests that the EDIT-B test performance is stable across the day. Finally, the study was prospective but cross-sectional, although based on patients with a proven BD or MDD trajectory; a longitudinal study with first episode major depression would provide the greatest evidence in favor of the predictive value of the test[30]. To this end, the generalizability to high-genetic-risk populations is currently being addressed in an ongoing prospective study (NCT06507787).

## CONCLUSION

In summary, this study provides additional evidence supporting the validity and clinical performance of the EDIT-B test as a potential complement to the objective diagnosis of bipolar disorder in depressed patients. These findings contribute to the advancement of precision psychiatry and suggest that such molecular biomarkers could help reduce misdiagnosis and the risk of inappropriate treatment in clinical practice.

## Data Availability

All data produced in the present study are available upon reasonable request to the authors

## Acknowledgements

The authors would like to acknowledge the contributions of Aixial for their support in conducting the study.

A first version of this manuscript has been made publicly available on the *medRxiv* preprint server (DOI: 10.1186/s12991-024-00544-8). The preprint has not undergone peer review and may differ from the final published version.

## Author contributions

EV, DW: lead the study, secured funding.

EV, DW, MF: conceived, designed the study

LVK, CH, JMH, EV: diagnosed patient, supervised and collected data.

DW, NS, FCR: draft the initial manuscript.

NS, FCR: prepared figures and tables and conducted data analyses and interpretation.

FSS, LVK, CH, JMH, EV: reviewed and edited the manuscript.

CC: conceived and designed biological protocols, provided support in conducting the study.

DV: clinical monitoring and managed the study.

LWB, GR: performed analyses.

AMM, VC, OSA, JZ, JZ, AGP, MV: diagnosed patient and collected data.

## Funding

This work was supported by EIT Health [220628/230125].

## Ethics approval and consent to participate

Ethical approval has been obtained in France by the research ethics committee Ile-de-France X (ID 2021-A-03194-37), in Spain by the research ethics committees of the Hospital Clinic of Barcelona (ID HCB/2022/0042) and the Sant Joan de Déu Foundation (PS-05-22) and in Denmark by the De Videnskabsetiske Komiteer for Region Hovedstaden (the Scientific Ethics Committees for the Capital Region of Denmark) (ID H-22010899).

## Disclosures/Competing interests

DW serves as Alcediag CSO.

NS, CC, FCR, DV are employed by Alcediag.

LWB, GR, MF are employed by Synlab.

EV has received grants and served as consultant, advisor or CME speaker for the following entities: AB-Biotics, Abbott, AbbVie, Adamed, Adium, Alcediag, Angelini, Biogen, Beckley-Psytech, Biohaven, Boehringer-Ingelheim, Casen-Recordati, Celon Pharma, Compass, Dainippon Sumitomo Pharma, Esteve, Ethypharm, Ferrer, Gedeon Richter, GH Research, Glaxo-Smith Kline, HMNC, Intra-Cellular therapies, Idorsia, Johnson & Johnson, Lundbeck, Luye Pharma, Medincell, Merck, Mitsubishi Tanabe Pharma, Newron, Novartis, Organon, Orion Corporation, Otsuka, Roche, Rovi, Sage, Sanofi-Aventis, Sunovion, Takeda, Teva, and Viatris, outside the submitted work.

LVK has within recent three years been a consultant for Teva and Lundbeck.

JZ has within recent three years received honoraria from Lundbeck.

AGP has received CME-related honoraria, or consulting fees from Janssen-Cilag, Casen Recordati and Rovi.

MV has received grants from Eli Lilly & Co.; and has served as a speaker for Abbott, Bristol-Myers Squibb, Glaxo-Smith Kline, Johnson & Johnson, and Lundbeck.

All other authors report no biomedical financial interests or potential conflicts of interest.

The described biomarker panel has been patented under the N° WO2021089866A1 and N° WO2024133568.

## Data availability statement

No data are available. Due to French, Danish and Spain legislation concerning data privacy, we are unable to share data from this study with others. Researchers wishing to access this data may submit a request to the relevant French, Danish, or Spanish authorities. Statistical analysis codes can be made available upon request.

## Notes

### Competing Interest Statement

DW is the CSO of Alcediag.
NS, CC, FCR, DV are employed by Alcediag.
LWB, GR, MF are employed by Synlab.
EV has received grants and served as consultant, advisor or CME speaker for the following entities: AB–Biotics, Abbott, AbbVie, Adamed, Adium, Alcediag, Angelini, Biogen, Beckley–Psytech, Biohaven, Boehringer-Ingelheim, Casen-Recordati, Celon Pharma, Compass, Dainippon Sumitomo Pharma, Esteve, Ethypharm, Ferrer, Gedeon Richter, GH Research, Glaxo-Smith Kline, HMNC, Intra–Cellular therapies, Idorsia, Johnson & Johnson, Lundbeck, Luye Pharma, Medincell, Merck, Mitsubishi Tanabe Pharma, Newron, Novartis, Organon, Orion Corporation, Otsuka, Roche, Rovi, Sage, Sanofi–Aventis, Sunovion, Takeda, Teva, and Viatris, outside the submitted work.
LVK has within recent three years been a consultant for Teva and Lundbeck.
CH, JMH has no conflict of interest.

### Clinical Trial

NCT05603819

### Clinical Protocols

https://doi.org/10.1186/s12991-024-00544-8

### Summary of Updates

This version represents the final manuscript accepted for publication in BMJ Mental Health (DOI: 10.1136/bmjment-2026-302859). Following a rigorous peer-review process, the manuscript has been profoundly revised, including revised figures and tables, revised supplementary files and additional analyses, particularly with regard to sensitivity analyses.

