## supplementary material for "EDIT-B Consortium: An External Pan-European Validation of an Innovative Machine Learning-Based Test for Bipolar Disorder Diagnosis"

#### Methods

#### Tables


- **Supplementary Table1.** EDIT-B study summary.
- **Supplementary Table2.** EDIT-B model hyper-parameters range and optimal parameters.
- **Supplementary Table3.** Demographic and clinical characteristics of patients included in the Analytical Set by clinical center.
- **Supplementary Table4.** Sociodemographic characteristics.
- **Supplementary Table5.** Clinical disease characteristics.
- **Supplementary Table6.** Mini-International Neuropsychiatric Interview (MINI).
- **Supplementary Table7.** Analysis of misclassified cases (False Positives and False Negatives) according to clinical and demographics covariates.
- **Supplementary Table8.** Diagnostic performances for EDIT-B test in analytical set and BD expert centers subgroup.
- **Supplementary Table9.** Summary of EDIT-B performance in stratified sensitivity analyses.


#### Figures


- **Supplementary Figure1.** Proposed neuroimmune mechanisms underlying the EDIT-B signature genes in mood disorders.
- **Supplementary Figure2.** STARD flow diagram of participants included in the EDIT-B consortium study.
- **Supplementary Figure3.** Precision-Recall curves plot of the EDIT-B test in analytical set and BD reference centers.
- **Supplementary Figure4.** ROC curves of EDIT-B test by BD reference center.
- **Supplementary Figure5.** Precision-Recall curves (PR-AUC) plot of the EDIT-B test by BD reference center.
- **Supplementary Figure6.** ROC curves analysis of the EDIT-B test in BD reference centers and comparison with the previous study.
- **Supplementary Figure7.** Target Editing Index analysis of EDIT-B targets.
- **Supplementary Figure8.** Density and calibration analysis plots of the EDIT-B model.
- **Supplementary Figure9.** Stratified sensitivity analysis according to disease characteristics.
- **Supplementary Figure10.** Stratified sensitivity analysis according to psychiatric comorbidities.
- **Supplementary Figure11.** Performance of the EDIT-B test across different demographics subtypes.
- **Supplementary Figure12.** Stratified sensitivity analysis according to young adult subpopulation.
- **Supplementary Figure13.** Stratified sensitivity analysis according to age of onset disease.
- **Supplementary Figure14.** Stratified sensitivity analysis according to illness duration.
- **Supplementary Figure15.** Stratified sensitivity analysis according to number of depression episode.
- **Supplementary Figure16.** EDIT-B True Positive (TP) and False Negative (FN) rates according to the number of manic, hypomanic, and mixed episodes.

- **Supplementary Figure17.** Stratified sensitivity analysis according to blood collection day-times.
- **Supplementary Figure18.** Correlation analysis between RNA editing biomarkers and clinical covariates.
- **Supplementary Figure19.** Decision Curve Analysis for the diagnostic performance of the EDIT-B model.

**Supplementary References**

**Supplementary Methods:**

### Eligibility criteria

Eligible participants were adults aged 18-80 years with a clinical diagnosis of BD or MDD, currently experiencing a major depressive episode (MDE). Diagnoses were confirmed according to DSM-5 criteria using the Mini-International Neuropsychiatric Interview (MINI)[1]. Both inpatients and outpatients were eligible. A Montgomery-Åsberg Depression Rating Scale (MADRS)[2] score of 20 or higher at assessment was required for inclusion. BD participants must have had at least one prior manic or hypomanic episode, while MDD participants must have had at least one previous MDE.
The exclusion criteria for participants are a Young Mania Rating Scale (YMRS)[3] of 12 or higher at the time of assessment, pregnancy, major central nervous system (CNS) affection, a diagnosis of schizoaffective disorder or having first-degree family history of BD for MDD participants.

### Sample size calculation

The sample size was determined to ensure adequate precision in estimating test performance metrics (including sensitivity, specificity, accuracy, and AUC-ROC) based on previously obtained results[4]. A total sample size of 392 patients with depression was calculated to estimate the performance parameters with a precision of ±5%, using the methods proposed by Buderer[5]. Considering a maximal 10% attrition rate, 436 patients were supposed to be included.

### Data collection and management

Data were recorded in an electronic case report form (e-CRF) by investigators and/or coordinators, with automated online checks for data integrity. After completion, final datasets were extracted, locked, and certified. The dataset was screened for missing values, and none were identified, confirming the completeness of the dataset. Participant data were pseudonymized and EDIT-B results were blinded to investigators until study completion (Supplementary Figure3).

### EDIT-B algorithm development

EDIT-B is a locked ML algorithm with a fixed function based on ExtraTree (ET)[6] method, one of the most recent machine learning algorithms for classification, using 22 RNA editing biomarkers, demographic data (age, sex), and psychiatric treatments (antidepressants, antiepileptics, antipsychotics (including lithium), anxiolytics, and hypnotics/sedatives, as well as alcohol and tobacco use. ExtraTrees method, derived from random Forest, adds another layer of randomness to decision forests: instead of searching for the optimal cut-point at each node during training, a random threshold is selected for each feature. ET forest requires the use of a training set used to construct the model and a test set to validate it. Development cohort (initial cohort used to develop and calibrate the EDIT-B algorithm prior to independent validation) was splitted into two samples: 60% of the dataset was used for the learning phase (n=147) and 40% was used for the testing phase (n=98). This splitting has been randomized and respected the initial proportion of the various status in each sample. We used a grid learning approach for each individual tree, where we stated certain maximum parameter sets (ntree= (500,1000), mtry= (1,20), splitrule=" extratrees" and min.node.size= (1,15)). The subtrees were trained with a classical k-fold cross-validation (k= (5,10)). EDIT-B model results were performed on the test dataset (N=98 samples)[4] that has never been used by the algorithm, the Switzerland multicentric study (N=143 samples)[4] and the European multicentric double-blind study (this study, N=393 samples). The implementation was done using the R ranger package (version 0.13.1)[7] and R caret package (version 6.0-90)[8]. The process was randomly repeated 500 times. To address the class imbalance between 63% MDD and 37% BD patients, various correction methods, including multi-downsizing techniques, were tested during the development phase. However, these were not selected for the final model as they did not provide a significant improvement in performance. After removing all the models with a CV lower than 15% in reproducibility and repeatability, for the probability scores, the final model was selected as the one that best minimized the training error and maximized the AUC ROC in the test dataset. The calibration of final model was performed on the development cohort. To ensure consistency across different datasets and centers, a standardized preprocessing pipeline (established during the development phase) was applied identically to all samples. This pipeline included Box-Cox normalization and z-score standardization. Because these preprocessing steps were fixed and applied uniformly, and because all samples were independently processed in a centralized external laboratory, no site-specific harmonization was required.

**Data analysis**

ROC curves and DeLong's test (pROC R package[9]) were used to evaluate binary classification performance and compare AUC-ROC values across populations. Precision-Recall curves plot the positive predictive value (precision) against the sensitivity (recall) at various threshold settings. The Area Under Precision-Recall (AUC-PR) summarizes this curve in a single value, representing the model's ability to correctly identify positive instances while minimizing false positives. These analysis were calculated using the precrec R package[10]. Model calibration and the distribution of predicted probabilities were assessed using a calibration plot (comparing predicted probabilities to the observed outcomes) and a density plot. A “Target Editing Index” (TEI), to summarize gene-specific RNA editing values, was calculated by linear combination of significant RNA editing variants maximizing AUC-ROC[11]. TEI values were centered-scaled and normalized using Box-Cox transformation for each gene in the development cohort and the same pre-processing step, parameters and formula was applied to the validation cohorts. Decision curve analysis was performed with the rmda R package[12] to assess the net benefit of the diagnostic test across a wide range of threshold probabilities, balancing the gain from correctly identifying bipolar disorder against the harm of misclassifying unipolar depression.

**Supplementary Table1. EDIT-B study summary**

| | |
|---|---|
| **Study title** | Clinical validation study for EDIT-B test: an aid for differential diagnosis of bipolar disorder, based on RNA editing blood biomarkers |
| **Short title** | EDIT-B |
| **Study participants** | In- and out adult patients between 18 and 80 years old diagnosed with bipolar disorder or major depressive disorder, and currently treated for an acute major depressive episode |
| **Study centers** | 4 centers in 3 countries (Denmark, France, Spain)<br>• Center1: Hospital Clinic de Barcelona (Spain)<br>• Center2: Psykiatrisk Center København (Denmark)<br>• Center3: GHU Paris Psychiatrie & Neurosciences (France)<br>• Center4: Parc Sanitari Sant Joan de Déu (Spain) |
| **Central laboratory** | SYNLAB Italia – Genetic Unit<br>via Beato L. Pavoni 18 25014 Castenedolo (BS) (Italy) |
| **Start date** | June 2022 |
| **Planned end date** | December 2024 |
| **Ethics approval and registration number** | France/BRC ID: 2021-A03062-39 |
| | Denmark/Journal-nr.: H-22010899 |
| | Spain (Hospital Clinic): Reg. HCB/2022/0042 |
| | Spain (Parc Sanitari Sant Joan de Déu): C.I. PS-05-22 |
| | ClinicalTrials.gov ID: NCT05603819 |
| **Study objectives and methods** | The objective of this study is to estimate three EDIT-B signatures in term of their external validity. For this purpose, performance of the test will be estimated by calculating for each signature its sensitivity, specificity and its accuracy to predict the diagnosis of bipolar disorder. Area under the ROC curve of each signature will be calculated. Additionally, positive predictive value, negative predictive value will be calculated. |

**Supplementary Table2. EDIT-B model hyper-parameters range and optimal parameters.** Hyper-parameters of grid search for the EDIT-B model using the caret package in R. Abbreviations: mtry= the number of variables randomly sampled as candidates at each split; min.node.size= the minimum number of observations that must be in a terminal node; ntree= the number of trees in the forest; train.control= the number of folds to use in k-fold cross-validation.

| **Parameters** | **Label** | **Searched Range** | **Optimal value** |
| --- | --- | --- | --- |
| mtry | Splitting Rule | [1,2,3,4,5,6,7,8,9,10,11,12,13,14,15,16,17,18,19,20] | 2 |
| min.node.size | Minimal Node Size | [1,2,3,4,5,6,7,8,9,10,11,12,13,14,15] | 1 |
| ntree | Number of trees | [500,1000] | 1000 |
| train.control | k-fold cross-validation | [5,10] | 10 |

**Supplementary Table3. Demographic and clinical characteristics of patients included in the Analytical Set by clinical center.** Data are the mean ± SEM. p-values of main characteristics are obtained with the student's t-test or chi-squared test. Effect size is calculated using [a]Cohen's d. Benchmarks: |d| ≈ 0.2 small, ≈ 0.5 medium, ≈ 0.8 large. [b]Odds ratio. [c]Cramér's V. Benchmarks: ≈ 0.1, small; ≈ 0.3, medium; ≈ 0.5, large. Abbreviations: MDD: Major depressive disorder patients; BD: bipolar disorder patients; MADRS: Montgomery-Åsberg Depression Rating Scale; YMRS: Young Mania Rating Scale; BMI: Body Mass Index.

| | Center 001 - BD reference center | | | | | Center 002 - BD reference center | | | | |
|---|---|---|---|---|---|---|---|---|---|---|
| | Total | MDD | BD | Effect Size | P-value | Total | MDD | BD | Effect Size | P-value |
| **Number, n** | 70 | 41 | 29 | | | 107 | 52 | 55 | | |
| **Age** | | | | | | | | | | |
| Age (min-max) | 24-79 | 24-73 | 24-79 | | | 18-78 | 19-78 | 18-76 | | |
| Age (mean ±SD) | 50.9±12.8 | 51.0±12.0 | 50.8±14.1 | 0.015[a] [-0.46, 0.49] | 0,9 | 42.0±15.6 | 44.9±15.4 | 39.2±15.5 | 0.37[a] [-0.01, 0.75] | 0,1 |
| **Sex** | | | | | | | | | | |
| Male (n (%)) | 26 (37.1) | 15 (36.6) | 11 (37.9) | 1.06[b] [0.40, 2.83] | 0,9 | 48 (44.9) | 22 (42.3) | 26 (47.3) | 1.22[b] [0.57, 2.62] | 0,6 |
| Female (n (%)) | 44 (62.9) | 26 (63.4) | 18 (62.1) | | | 59 (55.1) | 30 (57.7) | 29 (52.7) | | |
| **Race** | | | | | | | | | | |
| Asian | 0 (0) | 0 (0) | 0 (0) | | | 4 (3.7) | 2 (3.8) | 2 (3.6) | | |
| Black | 3 (4.3) | 1 (2.4) | 2 (6.9) | | | 5 (4.7) | 3 (5.8) | 2 (3.6) | | |
| White/Caucasian | 66 (94.3) | 39 (95.1) | 27 (93.1) | | | 98 (91.6) | 47 (90.4) | 51 (92.7) | | |
| Other | 1 (1.4) | 1 (2.4) | 0 | | | 0 | 0 | 0 | | |
| All | | | | 0.15[c] [0.00, 0.35] | 0,5 | | | | 0.05[c] [0.00, 0.18] | 0,5 |
| **BMI, kg/m²** (mean±SEM) | 27.3±5.1 | 27.0±5.7 | 27.7±4.4 | -0.14[a] [-0.62, 0.33] | 0,5 | 25.7±5.8 | 25.6±5.9 | 25.8±5.7 | -0.02[a] [-0.40, 0.35] | 0,9 |
| **Clinical Characteristics** | | | | | | | | | | |
| MADRS score (mean±SEM) | 33.5±6.1 | 33.7±5.5 | 33.1±6.9 | 0.01[a] [-0.37, 0.57] | 0,7 | 34.6±6.0 | 35.9±6.7 | 33.5±5.0 | 0.41[a] [0.02, 0.79] | <0.05 |
| YMRS score (mean±SEM) | 1.1±1.8 | 1.2±1.9 | 0.9±1.7 | 0.16[a] [-0.31 0.64] | 0,5 | 1.5±2.2 | 0.8±1.9 | 2.2±2.3 | -0.66[a] [-1.05, -0.27] | **<0.001** |
| **Psychotropic Treatments** | | | | | | | | | | |
| Anxiolytics (n (%)) | 27 (38.6) | 17 (41.5) | 10 (34.5) | 0.74[b] [0.28, 1.99] | 0,5 | 27 (25.2) | 20 (38.5) | 7 (12.7) | 0.23[b] [0.09, 0.62] | **<0.01** |
| Hypnotics/Sedatives (n (%)) | 9 (12.9) | 5 (12.2) | 4 (13.8) | 1.15[b] [0.28, 4.72] | 0,8 | 37 (34.6) | 27 (51.9) | 10 (18.2) | 0.21[b] [0.09, 0.49] | **<0.001** |
| Antiepileptics (n (%)) | 27 (38.6) | 6 (14.6) | 21 (72.4) | 15.31[b] [4.66, 50.27] | **<0.001** | 38 (35.5) | 6 (11.5) | 32 (58.2) | 10.67[b] [3.90, 29.15] | **<0.001** |
| Antipsychotics (n (%)) | 39 (55.7) | 14 (34.1) | 25 (86.2) | 12.05[b] [3.50, 41.54] | **<0.001** | 65 (60.7) | 19 (36.5) | 46 (83.6) | 8.88[b] [3.57, 22.06] | **<0.001** |
| Antidepressants (n (%)) | 61 (87.1) | 41 (100) | 20 (69) | 0.03[b] [0.00, 0.47] | **<0.001** | 60 (56.1) | 50 (96.2) | 10 (18.2) | 0.01[b] [0.00, 0.04] | **<0.001** |
| All treatement Classes | | | | 0.35[c] [0.16, 0.49] | 0,22 | | | | 0.55[c] [0.41, 0.67] | 0,24 |
| **Substance Addiction** | | | | | | | | | | |
| Tobacco (n (%)) | 22 (31.4) | 14 (34.1) | 8 (27.6) | | | 41 (38.3) | 14 (26.9) | 27 (49.1) | | |
| Alcohol (n (%)) | 3 (4.3) | 2 (4.9) | 1 (3.4) | | | 12 (11.2) | 6 (11.5) | 6 (10.9) | | |
| All | | | | 0.02[c] [0.00, 0.31] | 0,9 | | | | 0.14[c] [0.00, 0.41] | 0,9 |

| | Center 003 - BD reference center | | | | | Center 004 - general psychiatry center | | | | |
|---|---|---|---|---|---|---|---|---|---|---|
| | Total | MDD | BD | Effect Size | P-value | Total | MDD | BD | Effect Size | P-value |
| **Number, n** | 118 | 67 | 51 | | | 98 | 78 | 20 | | |
| **Age** | | | | | | | | | | |
| Age (min-max) | 18-76 | 18-75 | 19-76 | | | 20-79 | 20-79 | 28-73 | | |
| Age (mean ±SD) | 41.1±15.6 | 38.7±15.2 | 44.1±15.7 | -0.35[a] [-0.72, 0.02] | 0,1 | 53.6±12.3 | 53.1±12.4 | 55.3±12.1 | -0.18[a] [-0.67, 0.31] | 0,5 |
| **Sex** | | | | | | | | | | |
| Male (n (%)) | 42 (35.6) | 25 (37.3) | 17 (33.3) | 0.84[b] [0.39, 1.80] | 0,6 | 35 (35.7) | 29 (37.2) | 6 (30.0) | 0.72[b] [0.25, 2.09] | 0,5 |
| Female (n (%)) | 76 (64.4) | 42 (62.7) | 34 (66.7) | | | 63 (64.3) | 49 (62.8) | 14 (70.0) | | |
| **Race** | | | | | | | | | | |
| Asian | 1 (0.8) | 0 | 1 (2.0) | | | 1 (1.0) | 1 (1.3) | 0 | | |
| Black | 4 (3.4) | 2 (3.0) | 2 (3.9) | | | 1 (1.0) | 1 (1.3) | 0 | | |
| White/Caucasian | 100 (84.7) | 57 (85.1) | 43 (84.3) | | | 93 (94.9) | 73 (93.6) | 20 (100) | | |
| Other | 13 (11.0) | 8 (11.9) | 5 (9.8) | | | 3 (3.1) | 3 (3.8) | 0 | | |
| All | | | | 0.11[c] [0.00, 0.24] | 0,5 | | | | 0.12[c] [0.00, 0.25] | 0,7 |
| **BMI, kg/m²** (mean±SEM) | 25.8±5.5 | 26.2±5.7 | 25.2±5.2 | 0.18[a] [-0.19, 0.54] | 0,3 | 26.3±5.3 | 26.1±5.2 | 27.2±5.6 | -0.20[c] [0.69, 0.29] | 0,5 |
| **Clinical Characteristics** | | | | | | | | | | |
| MADRS score (mean±SEM) | 28.8±5.6 | 28.4±5.9 | 29.5±5.2 | -0.20[a] [-0.56, 0.17] | 0,3 | 32.4±6.7 | 31.8±5.8 | 34.4±9.1 | -0.39[a] [-0.89, 0.10] | 0,2 |
| YMRS score (mean±SEM) | 2.3±2.2 | 2.2±2.0 | 2.5±2.6 | -0.13[a] [-0.50, 0.23] | 0,5 | 0.5±1.1 | 0.4±0.9 | 1.2±1.5 | -0.76[a] [-1.27, -0.26] | **<0.05** |
| **Psychotropic Treatments** | | | | | | | | | | |
| Anxiolytics (n (%)) | 43 (36.4) | 25 (37.3) | 18 (35.3) | 0.92[b] [0.43, 1.96] | 0,8 | 42 (42.9) | 38 (48.7) | 4 (20.0) | 0.26[b] [0.08, 0.86] | **<0.05** |
| Hypnotics/Sedatives (n (%)) | 12 (10.2) | 5 (7.5) | 7 (13.7) | 1.97[b] [0.59, 6.62] | 0,3 | 25 (25.5) | 21 (26.9) | 4 (20.0) | 0.68[b] [0.20, 2.26] | 0,5 |
| Antiepileptics (n (%)) | 32 (27.1) | 5 (7.5) | 27 (52.9) | 13.95[b] [4.81, 40.44] | **<0.001** | 39 (39.8) | 24 (30.8) | 15 (75.0) | 6.75[b] [2.20, 20.70] | 0,15 |
| Antipsychotics (n (%)) | 80 (67.8) | 33 (49.3) | 47 (92.2) | 12.11[b] [3.92, 37.39] | **<0.001** | 65 (66.3) | 49 (62.8) | 16 (80.0) | 2.37[b] [0.72, 7.77] | 0,7 |
| Antidepressants (n (%)) | 93 (78.8) | 64 (95.5) | 29 (56.9) | 0.06[b] [0.02, 0.22] | **<0.001** | 95 (96.9) | 77 (98.7) | 18 (90.0) | 0.12[b] [0.01, 1.36] | **<0.05** |
| All treatement Classes | | | | 0.35[c] [0.21, 0.46] | 0,24 | | | | 0.21[c] [0.02, 0.30] | 0,24 |
| **Substance Addiction** | | | | | | | | | | |
| Tobacco (n (%)) | 36 (30.5) | 21 (31.3) | 15 (29.4) | | | 48 (49) | 39 (50) | 9 (45) | | |
| Alcohol (n (%)) | 8 (6.8) | 4 (6) | 4 (7.8) | | | 7 (7.1) | 7 (9) | 0 (0) | | |
| All | | | | 0.06[c] [0.00, 0.35] | 0,9 | | | | 0.17[c] [0.00, 0.43] | 0,2 |

**Supplementary Table4: Sociodemographic characteristics.** There were not significant differences in sociodemographic characteristics comparing MDD and BD patients. The majority of participants were single (42.2%) and lived either alone (36.6%) or with a partner (35.6%). Over half of the cohort reported a disability, predominantly temporary (37.7%). Educational levels were high, with 49.6% of patients holding a university degree and 41.5% having completed secondary school. Abbreviations: BD = bipolar disorder; MDD = major depressive disorder. P-value and effect-size were calculated using chi-squared test and Cramér's V method respectively.

| Social characteristics | All | MDD | BD | Effect Size (vs MDD) | P-value (vs MDD) |
|---|---|---|---|---|---|
| N | 393 | 238 | 155 | | |
| **Civil status** | | | | 0.05[a] [0.00, 0.12] | 0.9 |
| Single (n (%)) | 166 (42.2%) | 97 (40.8%) | 69 (44.5%) | | |
| Married (n (%)) | 126 (32.1%) | 77 (32.4%) | 49 (31.6%) | | |
| Separated/Divorced (n (%)) | 90 (22.9%) | 58 (24.4%) | 32 (20.6%) | | |
| Widowed (n (%)) | 11 (2.8%) | 6 (2.5%) | 5 (3.2%) | | |
| **Coexistence** | | | | 0.14[a] [0.00, 0.19] | 0.7 |
| Parents (n (%)) | 48 (12.2%) | 28 (11.8%) | 20 (12.9%) | | |
| Siblings (n (%)) | 21 (5.3%) | 16 (6.7%) | 5 (3.2%) | | |
| Children (n (%)) | 103 (26.2%) | 71 (29.8%) | 32 (20.6%) | | |
| Relatives (n (%)) | 4 (1.0%) | 4 (1.7%) | 0 | | |
| Spouse/Partner (n (%)) | 140 (35.6%) | 88 (37.0%) | 52 (33.5%) | | |
| Friends (n (%)) | 15 (3.8%) | 8 (3.4%) | 7 (4.5%) | | |
| Alone (n (%)) | 144 (36.6%) | 82 (34.5%) | 62 (40.0%) | | |
| Institution (n (%)) | 8 (2.0%) | 5 (2.1%) | 3 (1.9%) | | |
| **Current occupation** | | | | 0.17[a] [0.00, 0.28] | 0.2 |
| Unemployment (n (%)) | 71 (18.1%) | 43 (18.1%) | 28 (18.1%) | | |
| Temporary disability (n (%)) | 148 (37.7%) | 100 (42.0%) | 48 (31.0%) | | |
| Permanent disability (n (%)) | 55 (14.0%) | 36 (15.1%) | 19 (12.3%) | | |
| Retired (n (%)) | 52 (13.2%) | 30 (12.6%) | 22 (14.2%) | | |
| Active (n (%)) | 67 (17.0%) | 29 (12.2%) | 38 (24.5%) | | |
| **Academic level** | | | | 0.12[a] [0.00, 0.21] | 0.6 |
| No studies (n (%)) | 1 (0.3%) | 0 | 1 (0.6%) | | |
| Primary school (n (%)) | 33 (8.4%) | 22 (9.2%) | 11 (7.1%) | | |
| Secondary school (n (%)) | 163 (41.5%) | 106 (44.5%) | 57 (36.8%) | | |
| University degree (n (%)) | 195 (49.6%) | 109 (45.8%) | 86 (55.5%) | | |
| Unknown (n (%)) | 1 (0.3%) | 1 (0.4%) | 0 | | |

[a] Cramér's V. Benchmarks: ≈ 0.1, small; ≈ 0.3, medium; ≈ 0.5, large.

**Supplementary Table5: Clinical disease characteristics.** Compared with MDD, BD patients showed similar illness duration (~10–11 years) but more frequent (hypo)manic and depressive episodes, predominant depressive onset (88.4%), higher rates of psychotic symptoms and family history of affective and psychiatric disorders. Abbreviations: BD = bipolar disorder; MDD = major depressive disorder; SD = standard deviation. Psychiatric history: first and second generation. P-value and effect-size were calculated using Student's t-test or chi-squared test and Cohen's d or Odd Ratio (OR) method respectively.

| Disease characteristics | All | MDD | BD | Effect Size (vs MDD) | p-value (vs MDD) |
|---|---|---|---|---|---|
| **N** | 393 | 238 | 155 | | |
| **Diagnosis** | | | | | |
| BD I (n (%)) | 59 (15.0%) | - | 59 (38.1%) | | |
| BD II (n (%)) | 96 (24.4%) | - | 96 (61.9%) | | |
| BD (n (%)) | 155 (39.4%) | - | 155 (100%) | | |
| MDD (n (%)) | 238 (60.6%) | 238 (100%) | - | | |
| **Initial polarity** | | | | | |
| Depressive (n (%)) | | | 137 (88.4%) | | |
| Mixed (n (%)) | | | 2 (1.3%) | | |
| Manic/Hypomanic (n (%)) | | | 15 (9.7%) | | |
| UK (n (%)) | | | 1 (0.6%) | | |
| **Number of depression episodes** | | | | | |
| Mean (SD) | 5.7 (5.9) | 4.0 (4.3) | 8.7 (7.0) | -0.85[a] [-1.06, -0.64] | **<0.0001** |
| **Duration of illness (years)** | | | | | |
| Mean (SD) | 10.4 (13.1) | 10.1 (12.9) | 10.8 (13.4) | -0.05[a] [-0.26, 0.15] | 0.6 |
| **Age of onset of disease (years)** | | | | | |
| Mean (SD) | 29.9 (14.1) | 31.9 (14.7) | 26.7 (12.6) | 0.37[a] [0.17, 0.58] | **<0.001** |
| **Age of onset of the first depression (years)** | | | | | |
| Mean (SD) | 28.3 (14.0) | 31.3 (14.6) | 23.8 (11.7) | 0.55[a] [0.35, 0.76] | **<0.0001** |
| **Age of onset of the first mania (years)** | | | | | |
| Mean (SD) | — | — | 35.2 (16.2) | | |
| **Age of onset of the first hypomania (years)** | | | | | |
| Mean (SD) | — | — | 26.3 (10.6) | | |
| **Any history of affective pathology** | | | | | |
| No (n (%)) | 163 (41.5%) | 121 (50.8%) | 42 (27.1%) | | |
| Yes (n (%)) | 230 (58.5%) | 117 (49.2%) | 113 (72.9%) | 2.80[b] [1.50, 5.32] | **<0.001** |
| **Any history of psychotic symptoms** | | | | | |
| No (n (%)) | 326 (83.0%) | 222 (93.3%) | 104 (67.1%) | | |
| Yes (n (%)) | 67 (17.0%) | 16 (6.7%) | 51 (32.9%) | 6.48[b] [2.62, 18.44] | **<0.0001** |
| **Any family psychiatric history** | | | | | |
| No (n (%)) | 123 (31.3%) | 94 (39.5%) | 29 (18.7%) | | |
| Yes (n (%)) | 270 (68.7%) | 144 (60.5%) | 126 (81.3%) | 2.83[b] [1.44, 5.72] | **<0.005** |
| **Any family history of completed suicide** | | | | | |
| No (n (%)) | 309 (78.6%) | 191 (80.3%) | 118 (76.1%) | | |
| Yes (n (%)) | 76 (19.3%) | 43 (18.1%) | 33 (21.3%) | 1.23[b] [0.57, 2.65] | 0.5 |

[a] Cohen's d. Benchmarks (Cohen, 1988): |d| ≈ 0.2 small, ≈ 0.5 medium, ≈ 0.8 large.

[b] Odds ratio.

**Supplementary Table6: Mini-International Neuropsychiatric Interview (MINI).**

All participants had a current MDE with frequent past episodes, suicidal ideation and attempts were common in both groups, and psychiatric comorbidities (particularly panic disorder) were more frequent in BD. Abbreviations: MINI = Mini-International Neuropsychiatric Interview; BD = bipolar disorder; MDD = major depressive disorder; MDE = major depressive episode; PTSD = post-traumatic stress disorder.

| MINI | MDD | BD | BD I | BD II | All |
|---|---|---|---|---|---|
| **N** | 238 | 155 | 59 | 96 | 393 |
| **MDE - Time frame** | | | | | |
| Current (2 Weeks) (n (%)) | 238 (100%) | 155 (100%) | 59 (100%) | 96 (100%) | 393 (100%) |
| Past (n (%)) | 196 (82.4%) | 150 (96.8%) | 57 (96.6%) | 93 (96.9%) | 346 (88.0%) |
| Recurrent (n (%)) | 166 (69.7%) | 121 (78.1%) | 48 (81.4%) | 73 (76.0%) | 287 (73.0%) |
| **Manic episode** | | | | | |
| Yes (n (%)) | 0 | 62 (40.0%) | 59 (100%) | 3 (3.1%) | 62 (15.8%) |
| No (n (%)) | 238 (100%) | 93 (60.0%) | 0 | 93 (96.9%) | 331 (84.2%) |
| **Hypomanic episode** | | | | | |
| Yes (n (%)) | 0 | 99 (63.9%) | 5 (8.5%) | 94 (97.9%) | 99 (25.2%) |
| No (n (%)) | 238 (100%) | 56 (36.1%) | 54 (91.5%) | 2 (2.1%) | 294 (74.8%) |
| **Suicidality** | | | | | |
| Yes (n (%)) | 200 (84.4%) | 119 (76.8%) | 46 (78.0%) | 73 (76.0%) | 319 (81.4%) |
| No (n (%)) | 37 (15.6%) | 36 (23.2%) | 13 (22.0%) | 23 (24.0%) | 73 (18.6%) |
| **Suicide behavior disorder** | | | | | |
| Yes (n (%)) | 66 (27.8%) | 36 (23.2%) | 16 (27.1%) | 20 (20.8%) | 102 (26.0%) |
| No (n (%)) | 171 (72.2%) | 119 (76.8%) | 43 (72.9%) | 76 (79.2%) | 290 (74.0%) |
| **Any psychotic disorder** | | | | | |
| Yes (n (%)) | 0 | 15 (9.7%) | 13 (22.0%) | 2 (2.1%) | 15 (3.8%) |
| No (n (%)) | 238 (100%) | 140 (90.3%) | 46 (78.0%) | 94 (97.9%) | 378 (96.2%) |
| **BD with Psychotic Features** | | | | | |
| Yes (n (%)) | 0 | 27 (17.4%) | 27 (45.8%) | 0 | 27 (6.9%) |
| No (n (%)) | 238 (100%) | 128 (82.6%) | 32 (54.2%) | 96 (100%) | 366 (93.1%) |
| **Comorbidities** | | | | | |
| Agoraphobia (Yes, n (%)) | 21 (8.8%) | 15 (9.7%) | 5 (8.5%) | 10 (10.4%) | 36 (9.2%) |
| Any Psychotic Disorder (Yes, n (%)) | 0 | 15 (9.7%) | 13 (22.0%) | 2 (2.1%) | 15 (3.8%) |
| Antisocial Personality Disorder (Yes, n (%)) | 1 (0.4%) | 4 (2.6%) | 1 (1.7%) | 3 (3.1%) | 5 (1.3%) |
| Alcohol Use Disorder (Yes, n (%)) | 19 (8.0%) | 11 (7.1%) | 2 (3.4%) | 9 (9.4%) | 30 (7.6%) |
| Anorexia Nervosa (Yes, n (%)) | 0 | 0 | 0 | 0 | 0 |
| Binge-Eating Disorder (Yes, n (%)) | 3 (1.3%) | 5 (3.2%) | 0 | 5 (5.2%) | 8 (2.0%) |
| Bulimia Nervosa (Yes, n (%)) | 5 (2.1%) | 2 (1.3%) | 0 | 2 (2.1%) | 7 (1.8%) |
| Generalized Anxiety Disorder | 36 (15.1%) | 23 (14.8%) | 6 (10.2%) | 17 (17.7%) | 59 (15.0%) |
| Obessive-Compulsive Disorder (Yes, n (%)) | 23 (9.7%) | 10 (6.5%) | 3 (5.1%) | 7 (7.3%) | 33 (8.4%) |
| Panic disorder (Yes, n (%)) | 34 (14.3%) | 41 (26.5%) | 14 (23.7%) | 27 (28.1%) | 75 (19.1%) |
| PTSD (Yes, n (%)) | 17 (7.1%) | 6 (3.9%) | 0 | 6 (6.3%) | 23 (5.9%) |
| Social Anxiety Disorder (Yes, n (%)) | 22 (9.2%) | 19 (12.3%) | 4 (6.8%) | 15 (15.6%) | 41 (10.4%) |
| Substance Use Disorder (Yes, n (%)) | 10 (4.2%) | 7 (4.5%) | 3 (5.1%) | 4 (4.2%) | 17 (4.3%) |

**Supplementary Table7. Analysis of misclassified cases (False Positives and False Negatives) according to clinical and demographics covariates.** Detailed characterization of patients misclassified compared to correctly diagnosed individuals. Data are presented as means ±SD or percentages. P-values indicate whether a specific clinical feature significantly differentiates misclassifications from true classification. All p-values were adjusted for multiple comparison using the Benjamini-Hochberg method. Effect-size were calculated using odd ratio (OR) or mean difference (Δ mean) according to variable types. Only comorbidities with n>20 are included.

| Feature | N | Clinical MDD: True Negative | Clinical MDD: False Positive | Clinical MDD: Effect Size | Clinical MDD: P-value | Clinical BD: True Positive | Clinical BD: False Negative | Clinical BD: Effect Size | Clinical BD: P-value |
|---|---|---|---|---|---|---|---|---|---|
| MDD (n (%)) | 238 | 80.3% (191/238) | 19.7% (47/238) | — | — | — | — | — | — |
| BD (n (%)) | 155 | — | — | — | — | 82.6% (128/155) | 17.4% (27/155) | — | — |
| BD_I (n (%)) | 59 | — | — | — | — | 39.1% (50/128) | 33.3% (9/27) | OR 0.78 (0.33–Inf) | 1.0 |
| BD_II (n (%)) | 96 | — | — | — | — | 60.9% (78/128) | 66.7% (18/27) | OR 1.28 (0.57–Inf) | 1.0 |
| **Clinical Characteristics** | | | | | | | | | |
| Age (mean ±SD) | 393 | 46.7 (± 15.5) | 48.0 (± 12.8) | Δmean -1.34 (-5.67–2.99) | 0.7 | 45.6 (± 15.5) | 42.3 (± 17.4) | Δmean 3.30 (-4.04–10.64) | 1.0 |
| BMI (mean ±SD) | 381 | 26.0 (± 5.6) | 27.1 (± 5.5) | Δmean -1.14 (-3.05–0.77) | 0.5 | 26.5 (± 5.4) | 24.3 (± 4.6) | Δmean 2.19 (0.16–4.22) | 0.2 |
| MADRST (mean ±SD) | 393 | 32.2 (± 6.5) | 31.7 (± 6.7) | Δmean 0.46 (-1.71–2.63) | 0.8 | 32.0 (± 6.4) | 33.0 (± 6.3) | Δmean -1.02 (-3.74–1.70) | 1.0 |
| YMRST (mean ±SD) | 393 | 1.3 (± 1.9) | 0.5 (± 1.0) | Δmean 0.78 (0.38–1.18) | **<0.01** | 1.8 (± 2.2) | 2.6 (± 2.4) | Δmean -0.83 (-1.86–0.19) | 0.5 |
| Sex (n (%)) | 393 | 39.8% (76/191) | 31.9% (15/47) | OR 0.71 (0.33–1.46) | 0.6 | 37.5% (48/128) | 44.4% (12/27) | OR 1.33 (0.52–3.34) | 1.0 |
| **Race** | | | | | | | | | |
| Asian (n (%)) | 393 | 1.0% (2/191) | 2.1% (1/47) | OR 2.05 (0.03–40.11) | 0.6 | 2.3% (3/128) | 0.0% (0/27) | OR 0.00 (0.00–11.66) | 1.0 |
| Black (n (%)) | 393 | 3.7% (7/191) | 0.0% (0/47) | OR 0.00 (0.00–2.82) | 0.5 | 4.7% (6/128) | 0.0% (0/27) | OR 0.00 (0.00–4.08) | 1.0 |
| White (n (%)) | 393 | 89.0% (170/191) | 97.9% (46/47) | OR 5.66 (0.86–239.80) | 0.3 | 89.1% (114/128) | 100.0% (27/27) | OR Inf (0.73–Inf) | 0.6 |
| **Disease characteristics** | | | | | | | | | |
| Age of onset of disease in years (mean ±SD) | 391 | 31.3 (± 15.0) | 34.3 (± 13.5) | Δmean -3.02 (-7.48–1.45) | 0.5 | 27.2 (± 12.0) | 24.8 (± 15.2) | Δmean 2.37 (-3.95–8.70) | 1.0 |
| Age of onset of the first depression in years (mean ±SD) | 392 | 30.7 (± 14.7) | 33.6 (± 14.0) | Δmean -2.82 (-7.41–1.77) | 0.5 | 24.1 (± 11.2) | 22.6 (± 14.0) | Δmean 1.56 (-4.27–7.39) | 1.0 |
| Age of onset of the first hypomania in years (mean ±SD) | 100 | — | — | — | — | 26.4 (± 10.2) | 26.0 (± 12.7) | Δmean 0.38 (-6.07–6.83) | 1.0 |
| Age of onset of the first mania in years (mean ±SD) | 57 | — | — | — | — | 34.5 (± 15.6) | 39.1 (± 19.8) | Δmean -4.61 (-21.41–12.18) | 1.0 |
| Duration of illness in years (mean ±SD) | 392 | 9.9 (± 12.8) | 11.1 (± 13.5) | Δmean -1.19 (-5.52–3.15) | 0.7 | 10.9 (± 13.5) | 10.8 (± 13.1) | Δmean 0.06 (-5.60–5.72) | 1.0 |
| Number of depression episodes (mean ±SD) | 348 | 3.8 (± 4.1) | 4.9 (± 4.8) | Δmean -1.06 (-2.66–0.54) | 0.5 | 8.7 (± 7.1) | 8.2 (± 6.5) | Δmean 0.48 (-2.83–3.79) | 1.0 |
| Number of Hypomania episodes (mean ±SD) | 102 | — | — | — | — | 4.8 (± 5.7) | 4.1 (± 4.7) | Δmean 0.79 (-1.95–3.52) | 1.0 |
| Number of Mania episodes (mean ±SD) | 56 | — | — | — | — | 3.0 (± 2.8) | 0.9 (± 0.7) | Δmean 2.10 (1.16–3.04) | **<0.01** |
| Any history of psychotic symptoms (n (%)) | 393 | 5.8% (11/191) | 10.6% (5/47) | OR 1.94 (0.50–6.47) | 0.5 | 32.0% (41/128) | 37.0% (10/27) | OR 1.25 (0.47–3.19) | 1.0 |
| Any history of affective pathology (n (%)) | 393 | 52.4% (100/191) | 36.2% (17/47) | OR 0.52 (0.25–1.04) | 0.2 | 70.3% (90/128) | 85.2% (23/27) | OR 2.42 (0.75–10.26) | 0.6 |
| Any family psychiatric history (n (%)) | 393 | 63.9% (122/191) | 46.8% (22/47) | OR 0.50 (0.25–1.00) | 0.2 | 80.5% (103/128) | 85.2% (23/27) | OR 1.39 (0.42–6.04) | 1.0 |
| Any family history of completed suicide (n (%)) | 393 | 16.2% (31/191) | 25.5% (12/47) | OR 1.76 (0.75–3.97) | 0.4 | 21.9% (28/128) | 18.5% (5/27) | OR 0.81 (0.22–2.48) | 1.0 |
| Any psychotic disorder (n (%)) | 393 | — | — | — | — | 10.2% (13/128) | 7.4% (2/27) | OR 0.71 (0.07–3.45) | 1.0 |
| Psychotic features (n (%)) | 393 | — | — | — | — | 17.2% (22/128) | 18.5% (5/27) | OR 1.09 (0.29–3.42) | 1.0 |
| Suicidality (n (%)) | 392 | 85.3% (163/191) | 80.4% (37/46) | OR 0.71 (0.29–1.85) | 0.6 | 76.6% (98/128) | 77.8% (21/27) | OR 1.07 (0.37–3.55) | 1.0 |
| Suicide behavior disorder (n (%)) | 392 | 25.1% (48/191) | 39.1% (18/46) | OR 1.91 (0.91–3.95) | 0.2 | 23.4% (30/128) | 22.2% (6/27) | OR 0.93 (0.28–2.69) | 1.0 |
| **Comorbidities** | | | | | | | | | |
| Agoraphobia (n (%)) | 393 | 8.4% (16/191) | 10.6% (5/47) | OR 1.30 (0.35–3.99) | 0.7 | 10.2% (13/128) | 7.4% (2/27) | OR 0.71 (0.07–3.45) | 1.0 |
| Alcohol use disorder (n (%)) | 393 | 9.9% (19/191) | 0.0% (0/47) | OR 0.00 (0.00–0.82) | 0.1 | 3.1% (4/128) | 25.9% (7/27) | OR 10.58 (2.44–53.98) | **<0.01** |
| Generalised anxiety disorder (n (%)) | 393 | 13.6% (26/191) | 21.3% (10/47) | OR 1.71 (0.68–4.07) | 0.5 | 13.3% (17/128) | 22.2% (6/27) | OR 1.86 (0.54–5.71) | 0.8 |
| Obsessive-compulsive disorder (n (%)) | 393 | 9.9% (19/191) | 8.5% (4/47) | OR 0.84 (0.20–2.72) | 1.0 | 5.5% (7/128) | 11.1% (3/27) | OR 2.15 (0.33–10.27) | 1.0 |
| Panic Disorder (n (%)) | 392 | 15.7% (30/191) | 8.7% (4/46) | OR 0.51 (0.12–1.57) | 0.5 | 26.6% (34/128) | 25.9% (7/27) | OR 0.97 (0.32–2.66) | 1.0 |
| PTSD (n (%)) | 23 | 100.0% (14/14) | 100.0% (3/3) | OR 0.00 (0.00–Inf) | 1.0 | 100.0% (4/4) | 100.0% (2/2) | OR 0.00 (0.00–Inf) | 1.0 |
| Social Anxiety Disorder (n (%)) | 393 | 8.9% (17/191) | 10.6% (5/47) | OR 1.22 (0.33–3.70) | 0.8 | 12.5% (16/128) | 11.1% (3/27) | OR 0.88 (0.15–3.42) | 1.0 |

**Supplementary Table8. Diagnostic performances for EDIT-B test in analytical set and BD expert centers subgroup**. The BD reference center population demonstrated superior AUC-ROC (0.891, 95% CI: 0.854–0.929), as well as higher overall accuracy (84.1% vs. 81.2%), as compared to the full population (Analytical set). Abbreviations: AUC-ROC = Area under the receive operating characteristic curve; AUC-PR = Area under the Precision-Recall curve; CI95% = Confidence interval 95%; PPV: positive predictive value; NPV: negative predictive value, F1-score= Harmonic mean of PPV and Sensitivity, ECE= Expected Calibration Error.

| Diagnostic performances | Analytical set | BD reference centers |
|---|---|---|
| AUC-ROC [CI95%] | 0.873 [0.837 - 0.909] | 0.891 [0.854 - 0.929] |
| AUC-PR [CI95%] | 0.843 [0.798 - 0.884] | 0.892 [0.855 - 0.927] |
| Sensitivity (%) [CI95%] | 82.6% [75.7 - 88.2] | 81.5% [73.9 - 87.6] |
| Specificity (%) [CI95%] | 80.3% [74.6 - 85.1] | 86.3% [79.9 - 91.2] |
| PPV (%) [CI95%] | 73.1% [65.9 - 79.6] | 83.3% [75.9 - 89.3] |
| NPV (%) [CI95%] | 87.6% [82.5 - 91.7] | 84.7% [78.2 - 89.8] |
| Accuracy (%) [CI95%] | 81.2% [77.0 - 84.9] | 84.1% [79.4 - 88.1] |
| F1-score (%) [CI95%] | 77.6% [72.9 - 81.9] | 82.4% [77.6 - 86.9] |
| Odds ratio [CI95%] | 19.3 [11.4 - 31.7] | 27.6 [14.8 - 51.6] |
| Brier Score | 0.19 | 0.19 |
| ECE | 0.19 | 0.20 |
| True positives | 128 | 110 |
| True negatives | 191 | 138 |
| False positives | 47 | 22 |
| False negatives | 27 | 25 |
| N= | 393 | 295 |

**Supplementary Table9. Summary of EDIT-B performance in stratified sensitivity analyses.** Performance metrics of the EDIT-B algorithm across various stratified sensitivity on the analytical set. Reported values include sample size (N), AUC-ROC, overall accuracy (Acc), sensitivity (Se), and specificity (Sp). The table highlights the stability of the test's discriminative power regardless demographic and basic clinical characteristics, disease progression, clinical complexity, comorbidities and psychiatric treatment status. BD: bipolar disorder; MDD: major depressive disorder; MADRS: Montgomery-Åsberg Depression Rating Scale; AUC-ROC: area under the receiver operating characteristic curve.

| Variable | Groups | N | AUC-ROC | Acc (%) |
|---|---|---|---|---|
| Analytical Set | - | 393 | 0.873 | 81.2 |
| Bipolar reference centers | - | 295 | 0.891 | 84.1 |
| Sex | Male | 151 | 0.888 | 82.1 |
| | Female | 242 | 0.863 | 80.6 |
| Age Interval | [18-35] years | 118 | 0.877 | 80.5 |
| | [36-55] years | 157 | 0.894 | 80.3 |
| | >55 years | 118 | 0.865 | 83.1 |
| Young Subpopulation | [18-25] years | 47 | 0.902 | 85.1 |
| | [18-30] years | 85 | 0.917 | 84.7 |
| | [18-35] years | 118 | 0.877 | 80.5 |
| Bipolarity Subtype | BD I | 297 | 0.913 | 81.1 |
| | BD II | 334 | 0.849 | 80.5 |
| Onset Disease | <= 20 years | 132 | 0.874 | 81.1 |
| | > 20 years | 259 | 0.879 | 81.1 |
| | <= 26 years | 199 | 0.873 | 81.4 |
| | > 26 years | 192 | 0.884 | 80.7 |
| Illness Duration | <= 1 years | 143 | 0.893 | 85.3 |
| | <= 5 years | 211 | 0.864 | 82.0 |
| | >5 years | 181 | 0.885 | 78.5 |
| Depression Episodes | <=3 Episodes | 154 | 0.870 | 81.8 |
| | >3 Episodes | 194 | 0.849 | 82.0 |
| | <=4 Episodes | 209 | 0.881 | 83.3 |
| | >4 Episodes | 139 | 0.836 | 79.9 |
| MADRS Severity | Moderated | 251 | 0.878 | 80.5 |
| | Severe | 142 | 0.862 | 82.4 |
| Antidepressants | Treated | 309 | 0.839 | 79.3 |
| | Untreated | 84 | 0.868 | 88.1 |
| Antipsychotics | Treated | 249 | 0.851 | 76.3 |
| | Untreated | 144 | 0.823 | 89.6 |
| Anxiolytics | Treated | 139 | 0.913 | 87.1 |
| | Untreated | 254 | 0.846 | 78.0 |
| Hypnotics and Sedatives | Treated | 83 | 0.841 | 81.9 |
| | Untreated | 310 | 0.877 | 81.0 |
| Antiepileptics | Treated | 136 | 0.737 | 72.8 |
| | Untreated | 257 | 0.853 | 85.6 |
| Psychotic Symptoms | No | 326 | 0.875 | 81.9 |
| | Yes | 67 | 0.803 | 77.6 |
| Agoraphobia | No | 357 | 0.873 | 81.2 |
| | Yes | 36 | 0.870 | 80.6 |
| Alcohol Use Disorder | No | 363 | 0.886 | 81.5 |
| | Yes | 30 | 0.842 | 76.7 |
| Generalized Anxiety Disorder | No | 334 | 0.884 | 82.6 |
| | Yes | 59 | 0.815 | 72.9 |
| Obsessive-Compulsive Disorder | No | 360 | 0.879 | 81.4 |
| | Yes | 33 | 0.783 | 78.8 |
| Panic Disorder | No | 317 | 0.871 | 80.4 |
| | Yes | 75 | 0.908 | 85.3 |
| Social Anxiety Disorder | No | 352 | 0.874 | 81.3 |
| | Yes | 41 | 0.866 | 80.5 |
| 1st Degree Family History | No | 159 | 0.848 | 76.1 |
| | Yes | 234 | 0.899 | 84.2 |
| History of Affective Pathology | No | 163 | 0.883 | 79.1 |
| | Yes | 230 | 0.883 | 82.6 |
| Family Suicide History | No | 309 | 0.869 | 81.6 |
| | Yes | 76 | 0.863 | 77.6 |
| Family Psychiatry History | No | 123 | 0.869 | 76.4 |
| | Yes | 270 | 0.890 | 83.3 |
| Sample Collection Time | Before 11hours | 150 | 0.910 | 85.3 |
| | After 11hours | 145 | 0.871 | 82.8 |

**Supplementary Figure1. Proposed neuroimmune mechanisms underlying the EDIT-B signature genes in mood disorders.**

This figure illustrates a hypothetical mechanistic framework integrating the biological functions of genes included in the EDIT-B signature. (1) Type I interferon (IFN-I) and IL-17 signaling, mediated by IFNAR1 and IL17RA, respectively, activate transcriptional programs involved in immune responses and pro-inflammatory signaling through the JAK–STAT, ACT1/TRAF6, MAPK and NF-κB pathways[13–16]. ZNF267 is hypothesized to regulate transcriptional programs associated with inflammatory responses and mood disorders[17]. (2) PTPRC/CD45 regulates the activation state of the Src-family kinase LYN, a key signaling hub controlling immune-cell activation[18]. (3) Activated LYN promotes recruitment and phosphorylation of the adaptor protein GAB2[19], leading to activation of the PI3K–AKT pathway downstream of multiple immune receptors, including cytokine receptors, Fcγ receptors and TREM2-associated signaling[20]. (4) In parallel, PRKCB activates NF-κB/MAPK signaling, enhancing the production of pro-inflammatory mediators and inflammasome activation[21,22]. AKT-dependent regulation of MDM2 and the p53 pathway may further contribute to oxidative stress, mitochondrial dysfunction and amplification of inflammatory responses[23,24]. (5) Together, these converging pathways may promote sustained microglial activation, increased production of pro-inflammatory mediators, inflammasome activation, oxidative stress and aberrant synaptic pruning, resulting in chronic neuroinflammation and increased vulnerability to psychiatric disorders. Notably, experimental evidence demonstrates that direct IFNAR1 signaling in microglia induces a type I interferon gene signature accompanied by increased PTPRC/CD45 expression, complement (C4b) activation and enhanced synaptic engulfment, supporting a mechanistic link between peripheral immune activation and neurobiological dysfunction[25]. The proposed interactions represent a synthesis of current evidence and should be considered a biologically plausible working model rather than a demonstrated signaling cascade. Created with BioRender.com.

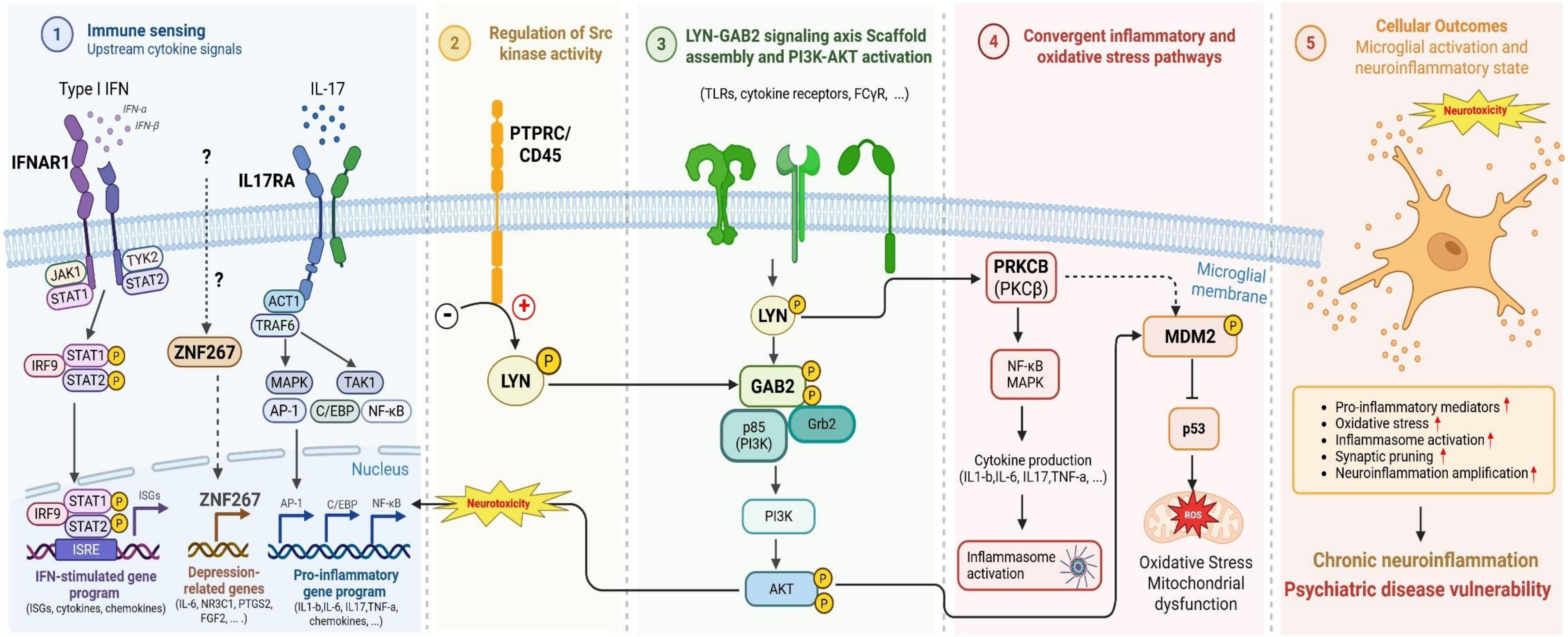
1 Immune sensing
Upstream cytokine signals
Type I IFN
IFN-α
IFN-β
IFNAR1
JAK1
STAT1
TYK2
STAT2
IL-17
IL17RA
ACT1
TRAF6
MAPK
TAK1
AP-1
C/EBP
NF-κB
?
?
ZNF267
IRF9
STAT1
STAT2
P
Nucleus
ISGs
ISRE
IFN-stimulated gene program
(ISGs, cytokines, chemokines)
ZNF267
Depression-related genes
(IL-6, NR3C1, PTGS2, FGF2, ... .)
AP-1
C/EBP
NF-κB
Pro-inflammatory gene program
(IL1-b,IL-6, IL17,TNF-a, chemokines, ...)
2 Regulation of Src kinase activity
PTPRC/ CD45
-
+
LYN
Neurotoxicity
3 LYN-GAB2 signaling axis Scaffold assembly and PI3K-AKT activation
(TLRs, cytokine receptors, FCγR, ...)
LYN
GAB2
Grb2
p85 (PI3K)
PI3K
AKT
4 Convergent inflammatory and oxidative stress pathways
PRKCB (PKCβ)
NF-κB MAPK
Cytokine production
(IL1-b,IL-6, IL17,TNF-a, ...)
Inflammasome activation
Microglial membrane
MDM2
p53
ROS
Oxidative Stress
Mitochondrial dysfunction
5 Cellular Outcomes
Microglial activation and neuroinflammatory state
Neurotoxicity
Pro-inflammatory mediators
Oxidative stress
Inflammasome activation
Synaptic pruning
Neuroinflammation amplification
Chronic neuroinflammation
Psychiatric disease vulnerability

**Supplementary Figure2. STARD flow diagram of participants included in the EDIT-B consortium study.** The EDIT-B consortium study is a multicenter study recruiting participants in four different centers in Spain, France and Denmark, respectively. Participants are assessed for eligibility with several inclusion and exclusion criteria during the inclusion visit. A total of 426 participants were screened for eligibility: 259 with MDD, 165 with BD, and two with missing diagnostic data. Eight participants did not meet the inclusion/exclusion criteria. From the other 418 individuals, 14 were excluded for protocol deviations (three missing clinician signatures on consent forms at database lock, one with family psychiatric history, one with central nervous system (CNS) disease and nine with improperly handled blood system) and 14 additional participants were excluded for intended use or technical quality criteria. The final analytical set included 393 participants: 238 with MDD and 155 with BD (3 participants were re-included in the analytical set following the delayed receipt of the Informed Consent Forms signed by the treating physicians).

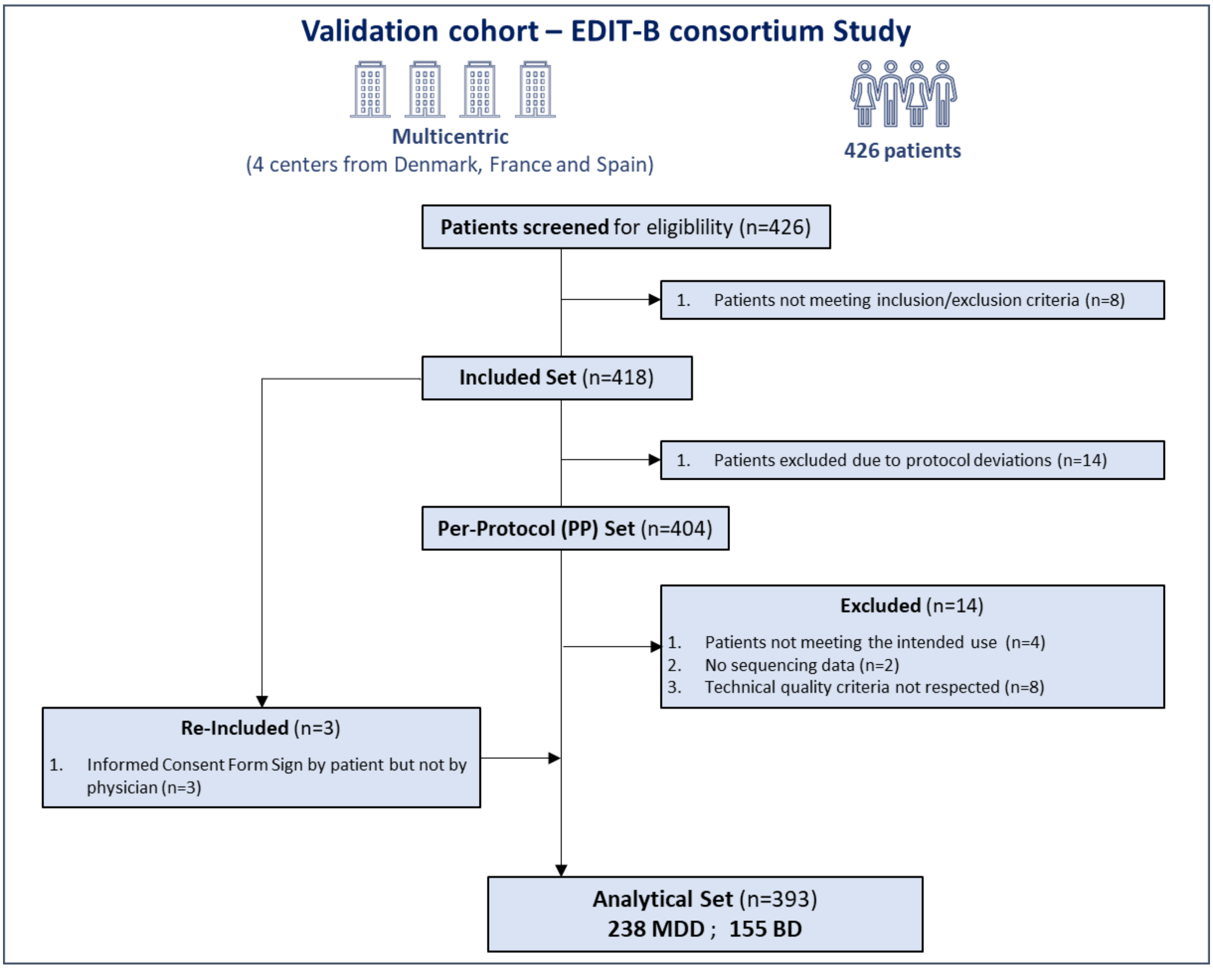

**Supplementary Figure3. Precision-Recall curves plot of the EDIT-B test in analytical set and BD reference centers.** A) AUC-PR curve for the analytical set performed in the present study (AUC-PR=0.843, N=393); B) AUC-PR curve for the BD reference centers set performed in the present study (AUC-PR=0.892, N=295); Coral: Optimal Recall–Precision values using the same threshold for the two cohorts; Dark blue: PR-AUC curves.

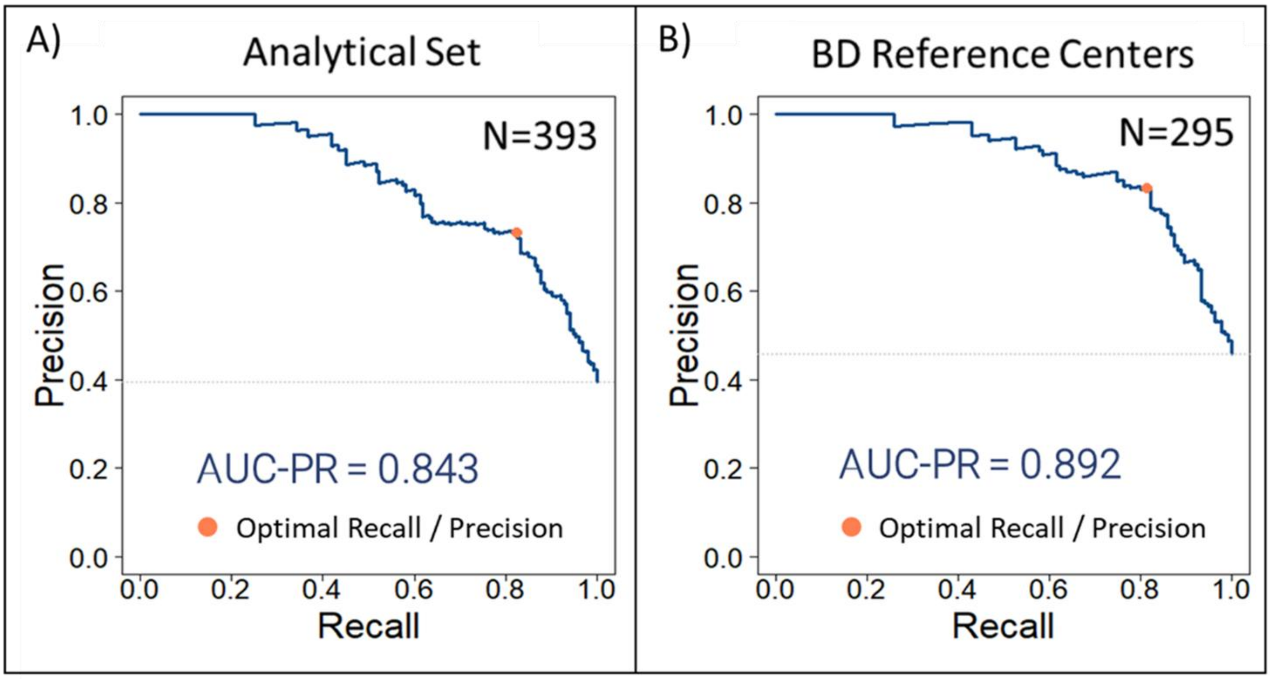

**Supplementary Figure4. ROC curves of EDIT-B test by BD reference center.**

An exploratory analysis was conducted to assess the AUC-ROC curves of EDIT-B test across the three BD reference centers. Overall, EDIT-B test demonstrated high performances across all three centers: A) BD reference center 001 (AUC=0.898, N=70); B) BD reference center 002 (AUC=0.887, N=107); C) BD reference center 003 (AUC=0.893, N=118). Dark blue line: Empirical ROC curve, Dotted light blue line: Generalized ROC curve.

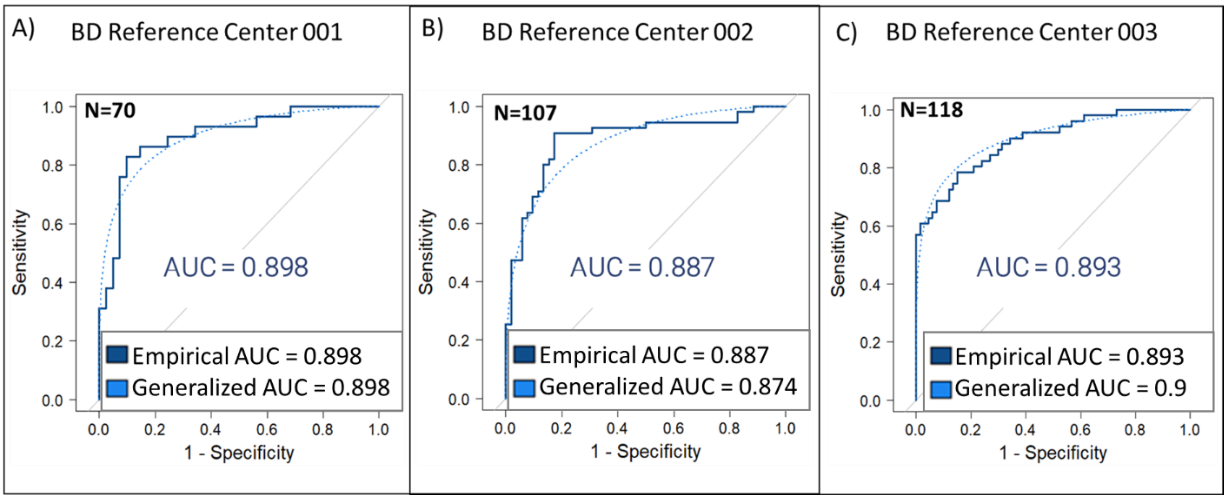

**Supplementary Figure5. Precision-Recall curves (PR-AUC) plot of the EDIT-B test by BD reference center.** A) BD reference center 001 (PR-AUC=0.898, N=70); B) BD reference center 002 (PR-AUC=0.887, N=107); C) BD reference center 003 (PR-AUC=0.893, N=118); Coral: Optimal recall-precision values using the same threshold for the three centers; Dark blue: PR-AUC curves.

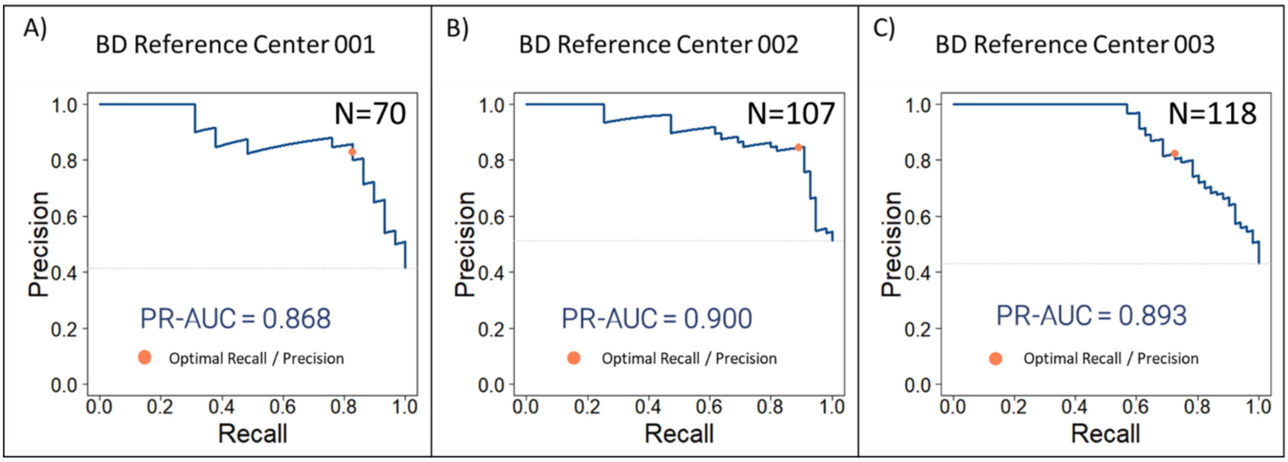

**Supplementary Figure6. ROC curves analysis of the EDIT-B test in BD reference centers and comparison with the previous study.**

A) Empirical and Generalized ROC curves of EDIT-B test in BD reference centers (AUC=0.891, N=295). Dark blue: Empirical ROC Curve, light blue: Generalized ROC Curve; B) ROC curve and confidence intervals comparison of the EDIT-B test in present bipolar reference centers vs previous study[25] (p=0.7). Coral: previous study; Dark blue: present study in BD reference centers; P-value = p-value of the DeLong's Test.

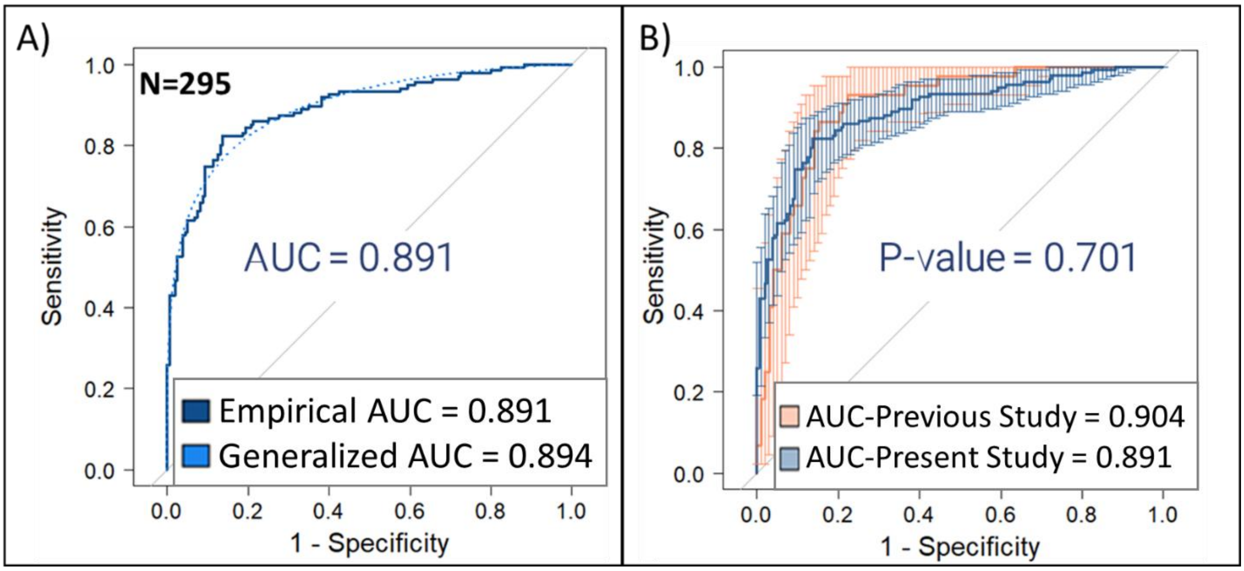

**Supplementary Figure7. Targeting Editing Index (TEI) analysis of EDIT-B targets.**

Barplot representations of the TEI values for EDIT-B genes across four distinct cohorts: (A) the development cohort (N=245) from Montpellier University Hospital, (B) the first validation cohort (N=143) from the Swiss study at Les Toises centers, (C) the second validation cohort (N=393) from the EIT-Health study involving four European centers, and (D) the combined first and second validation cohorts (N=536). Blue bars represent unipolar patients and coral bars represent bipolar patients. Significant differences between the two groups are indicated by * (* adjusted $p < 0.05$, ** adjusted $p < 0.01$, *** adjusted $p < 0.001$). TEI values were centered, scaled, and normalized using Box-Cox transformation for each gene in the development cohort, and the same pre-processing steps, parameters and formula were applied to the validation cohorts.

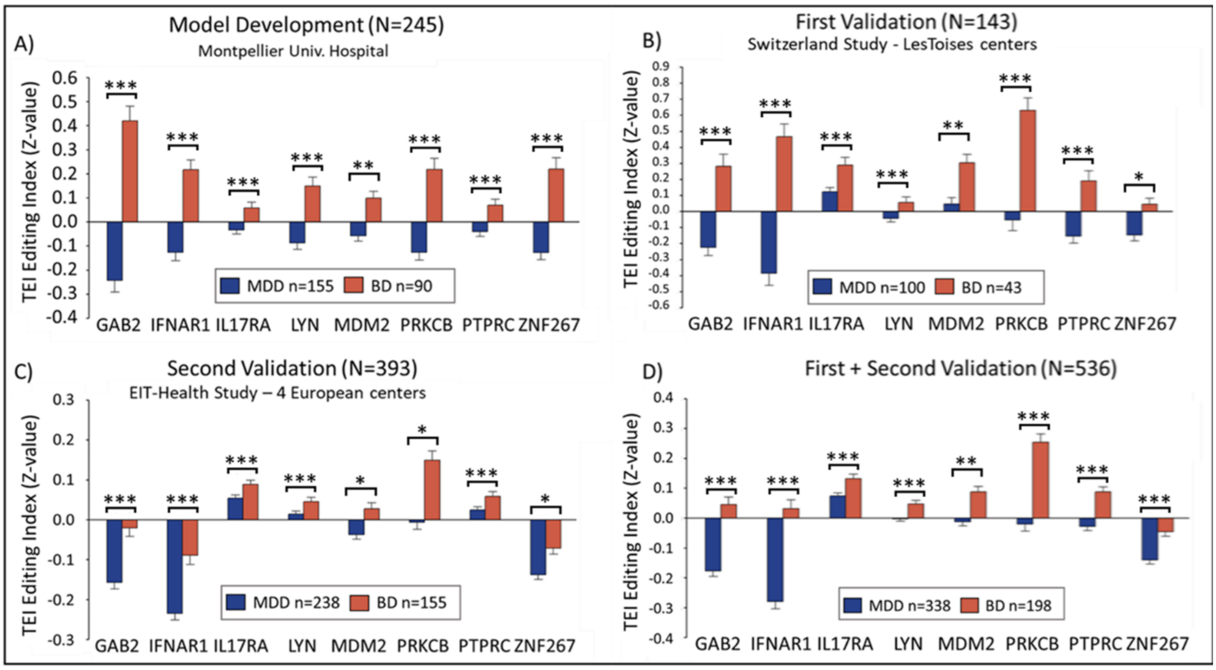

**Supplementary Figure8. Density and calibration analysis plots of the EDIT-B model.**

A) Density plot: The distribution of predicted scores shows both MDD and BD populations separated by the algorithm threshold. Light blue: MDD patients; Coral: BD patients; B) Calibration plot. A calibration plot comparing the observed proportion of positives outcomes vs the predicted probability. A low Brier score value indicates that the selected model is accurate and performs well in predicting the probabilities of the positive class. ECE: Expected Calibration Error. Dark blue points: patients' values, dark blue line: predicted calibration line, light blue: 95% quantile interval.

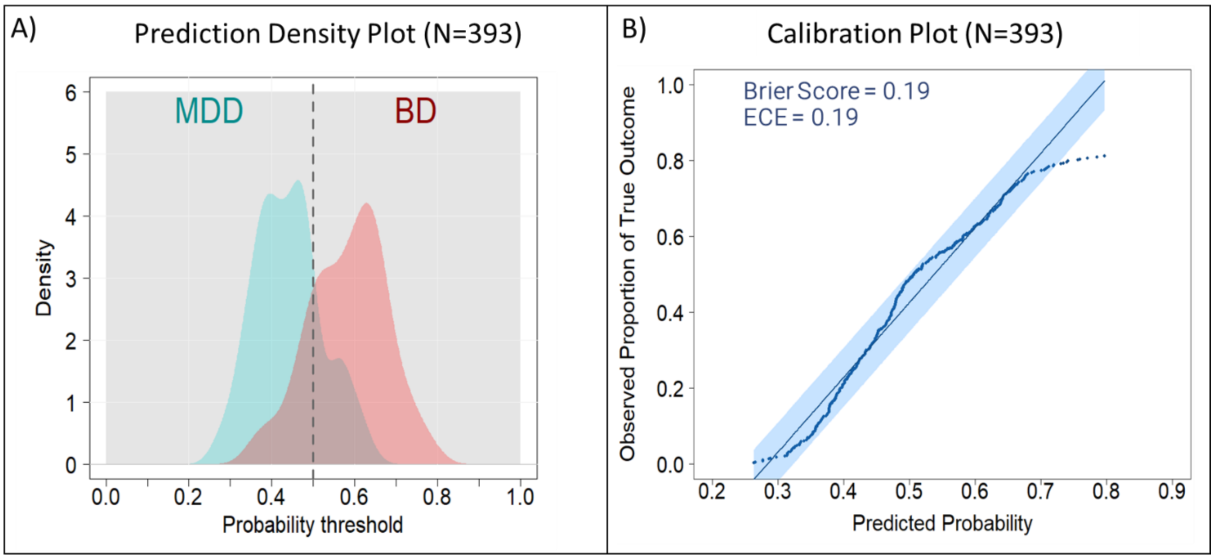

**Supplementary Figure9. Stratified sensitivity analysis according to disease characteristics.** Receiver operating characteristic (ROC) curves comparing the diagnostic performance of the EDIT-B test across several clinical and family history: (A) presence of psychotic symptoms, (B) first-degree family history, (C) history of affective pathology, (D) family history of completed suicide, (E) general family psychiatric history, and (F) depression severity based on MADRS scores (moderately vs. severely depressed). For all analyzed characteristics, no significant difference in the area under the curve (AUC) was observed between subgroups (all $p > 0.19$), indicating that the discriminative power of the EDIT-B test is robust and independent of these clinical features.

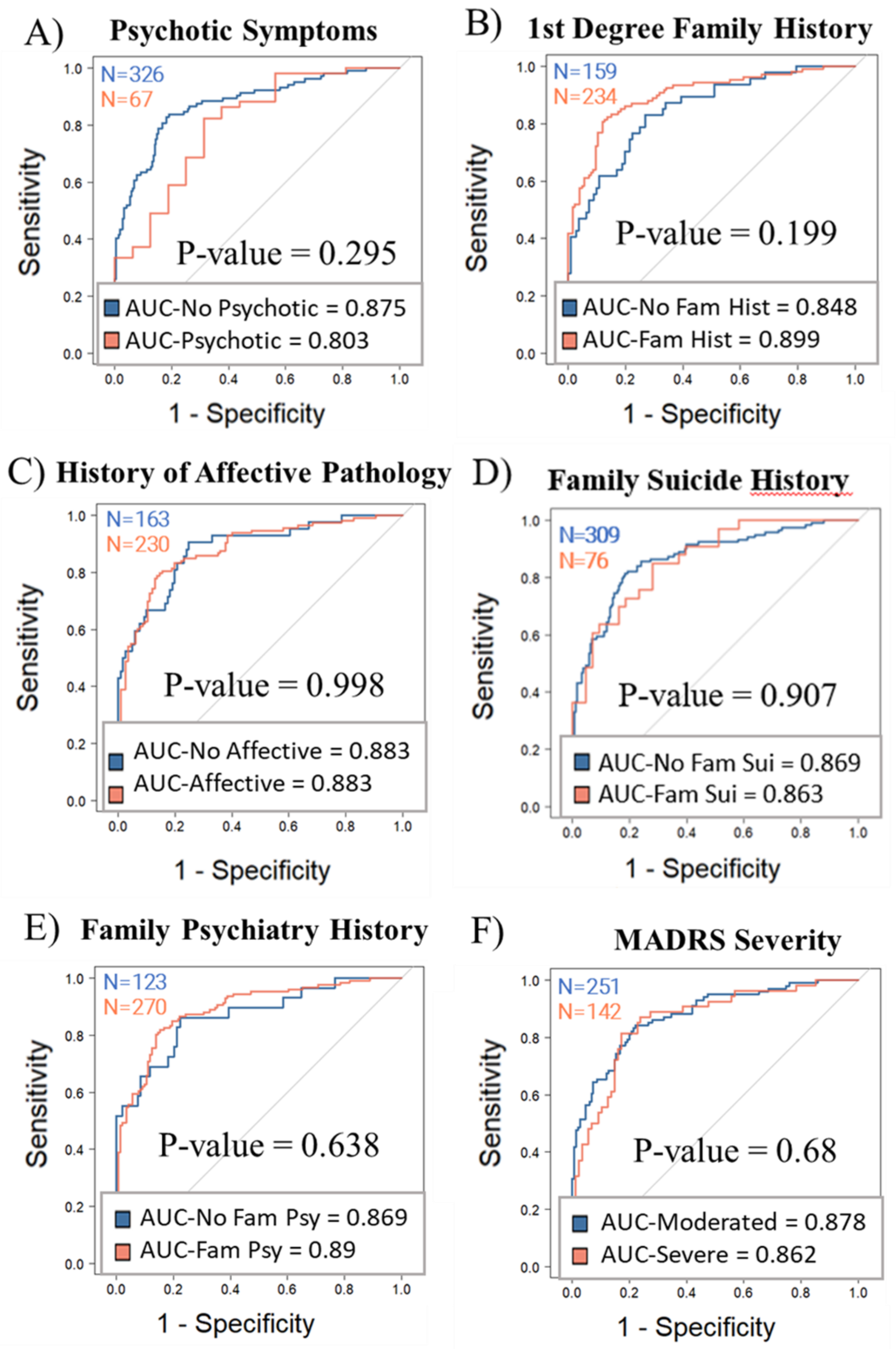

**Supplementary Figure10. Stratified sensitivity analysis according to psychiatric comorbidities.** Receiver operating characteristic (ROC) curves comparing the diagnostic performance of the EDIT-B test in patients with and without the following psychiatric comorbidities: **(A)** Agoraphobia, **(B)** Alcohol Use Disorder, **(C)** Generalized Anxiety Disorder, **(D)** Obsessive-Compulsive Disorder, **(E)** Panic Disorder, and **(F)** Social Anxiety Disorder. For all analyzed comorbidities, no significant difference in the area under the curve (AUC) was observed between the affected and unaffected populations (all $p > 0.25$). These results demonstrate that the discriminative power of the EDIT-B test is robust and independent of common psychiatric comorbidities.

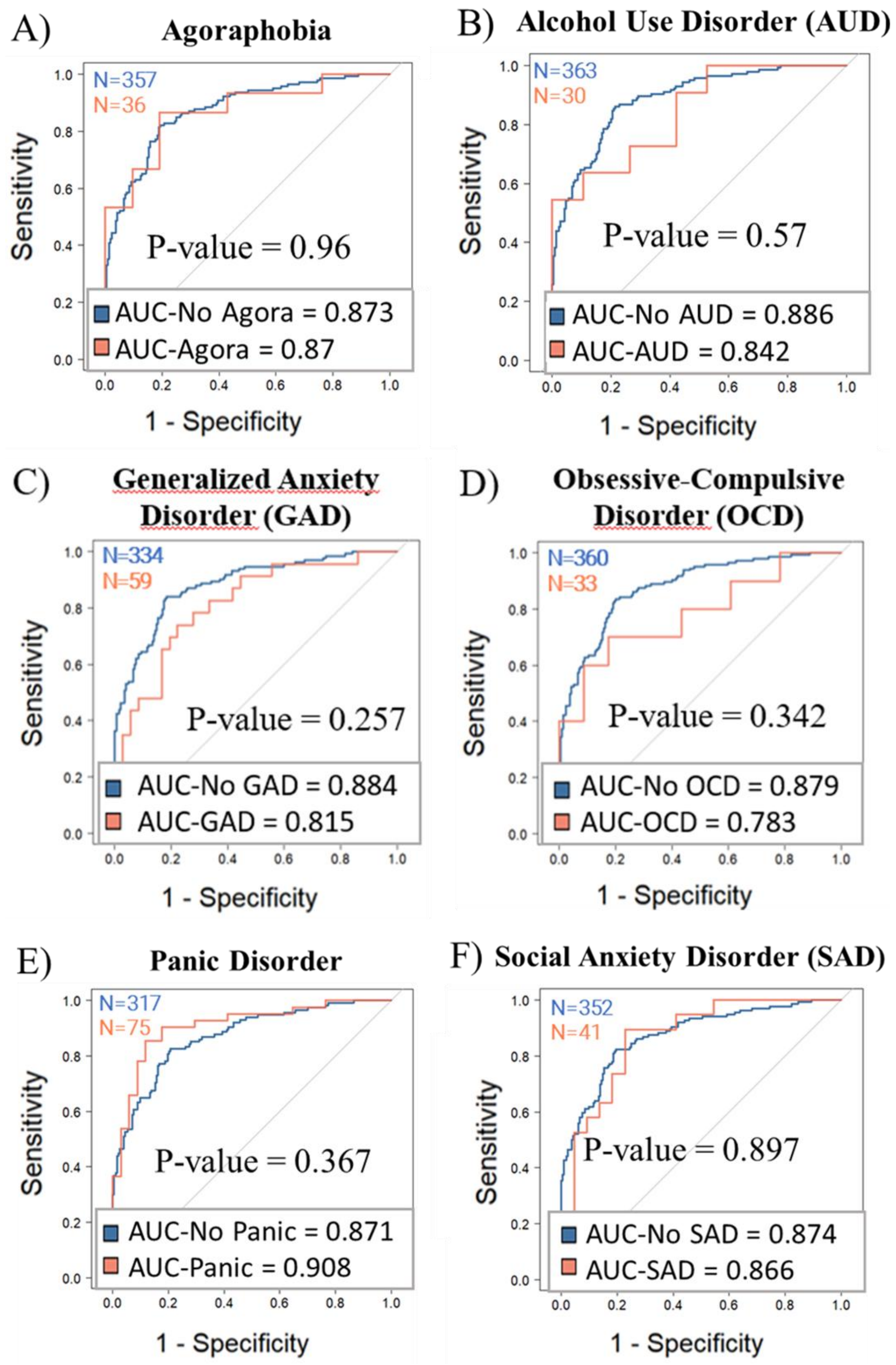

**Supplementary Figure11. Performance of the EDIT-B test across different demographics subtypes.** Receive Operating Characteristic (ROC) curves were generated to evaluate the diagnostic performance of EDIT-B test across sex, age and bipolar disorder subtype. A) The EDIT-B test by sex shows similar performance for males (AUC = 0.888) and females (AUC = 0.863) with a P-value of 0.49, indicating no significant difference. (B) The EDIT-B test across three age intervals ([18-35 years], [36-55 years], and [>55 years]) demonstrates consistent performance with AUC values of 0.877, 0.894, and 0.865, respectively, and a P-value of 0.92, indicating no significant difference among age groups. (C) The EDIT-B test by bipolar disorder (BD) subtype shows a statistically significant difference between BD-I (AUC = 0.913) and BD-II (AUC = 0.849) with a P-value of 0.04.

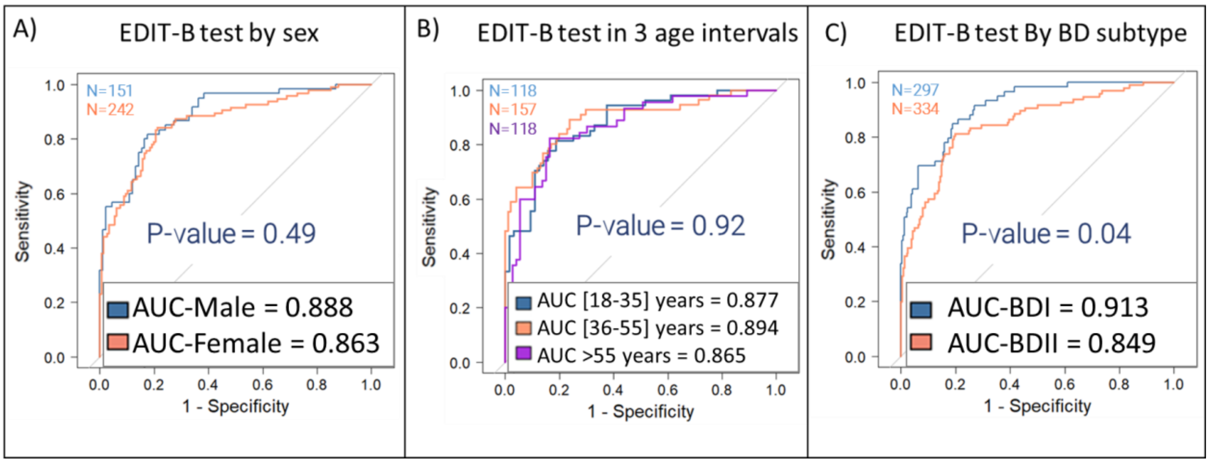

**Supplementary Figure12. Stratified sensitivity analysis according to young subpopulation.** (A) Age distribution histograms and density plots for the analytical set (N=393), patients with major depressive disorder (MDD, N=238), and patients with bipolar disorder (BD, N=155). The green shaded area highlights the younger subpopulation (up to 35 years), demonstrating a broad and representative age distribution across both diagnostic groups. (B) Receiver operating characteristic (ROC) curves comparing the diagnostic performance of EDIT-B across three young adult strata: 18–25 years (blue; AUC = 0.902, N=46, 24MDD and 22BD), 18–30 years (orange; AUC = 0.917, N=78, 41MDD and 37BD), and 18–35 years (purple; AUC = 0.877, N=109, 57MDD and 52 BD). No significant differences in area under the curve (AUC) were observed between these subgroups (p = 0.79, 0.66, and 0.36, respectively), confirming that the diagnostic accuracy of the EDIT-B test is maintained in young adult populations.

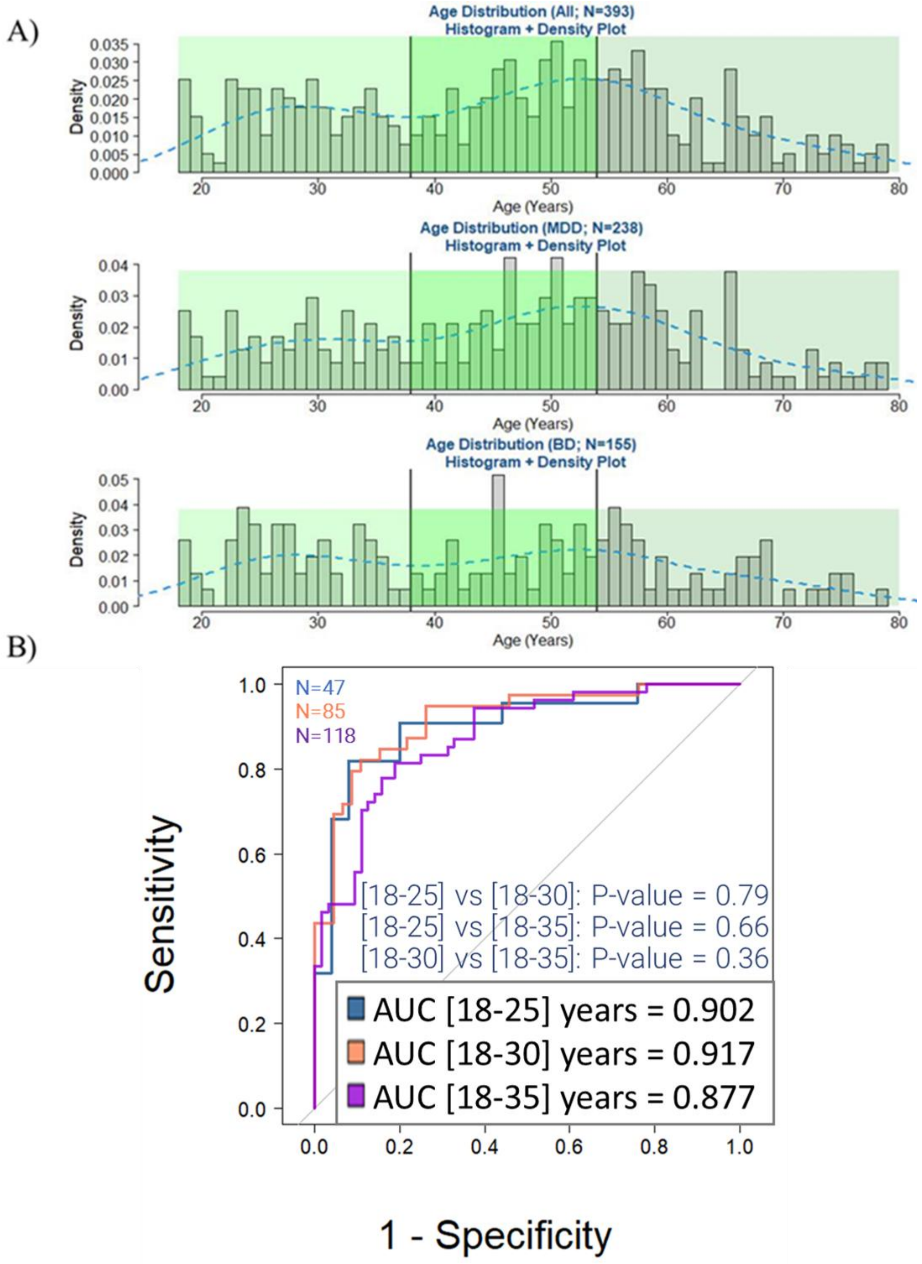

**Supplementary Figure13. Stratified sensitivity analysis according to age of onset disease.** (A) Density plot illustrating the distribution of the age of disease onset in the study population, highlighting the peak (maximum density) at 20 years and the median age of onset at 26 years. (B) Receiver operating characteristic (ROC) curves comparing the diagnostic performance of EDIT-B for patients with an age of onset ≤ 20 years (blue; AUC = 0.874) versus those with an age of onset >20 years (coral; AUC = 0.879). (C) ROC curves comparing performance for patients with an age of onset ≤ 26 years (blue; AUC = 0.873) versus those with an age of onset >26 years (coral; AUC = 0.884). No significant differences in area under the curve (AUC) were observed between these subgroups (p = 0.90 and p = 0.76, respectively), indicating that the discriminative power of the EDIT-B test is independent of the age at which the disease first manifested.

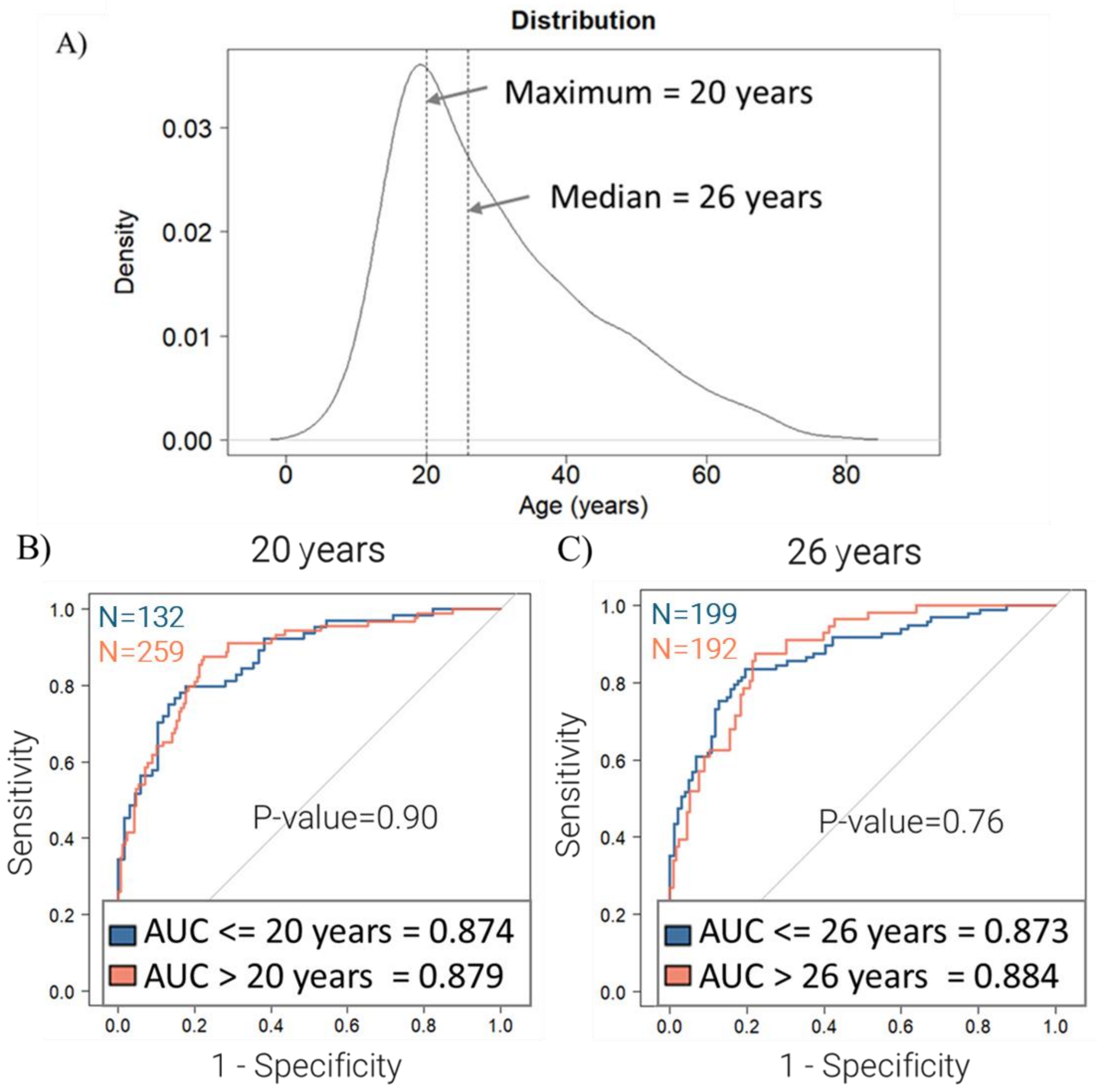

**Supplementary Figure14. Stratified sensitivity analysis according to illness duration.** Density plot showing the distribution of illness duration in the analytical set (n=393), with markers indicating the 1-year and 5-year thresholds. (B) Receiver operating characteristic (ROC) curves comparing the diagnostic performance of EDIT-B for three subgroups: illness duration ≤ 1 year (blue; AUC = 0.893; N=143, 79MDD vs 64BD), ≤ 5 years (orange; AUC = 0.864; N=211, 129MDD vs 82BD), and >5 years (purple; AUC = 0.885; N=181, 109MDD vs 72BD). No significant differences in area under the curve (AUC) were observed between these groups (all $p > 0.4$), indicating that the discriminative power of the EDIT-B test remains consistent regardless of the time elapsed since disease onset.

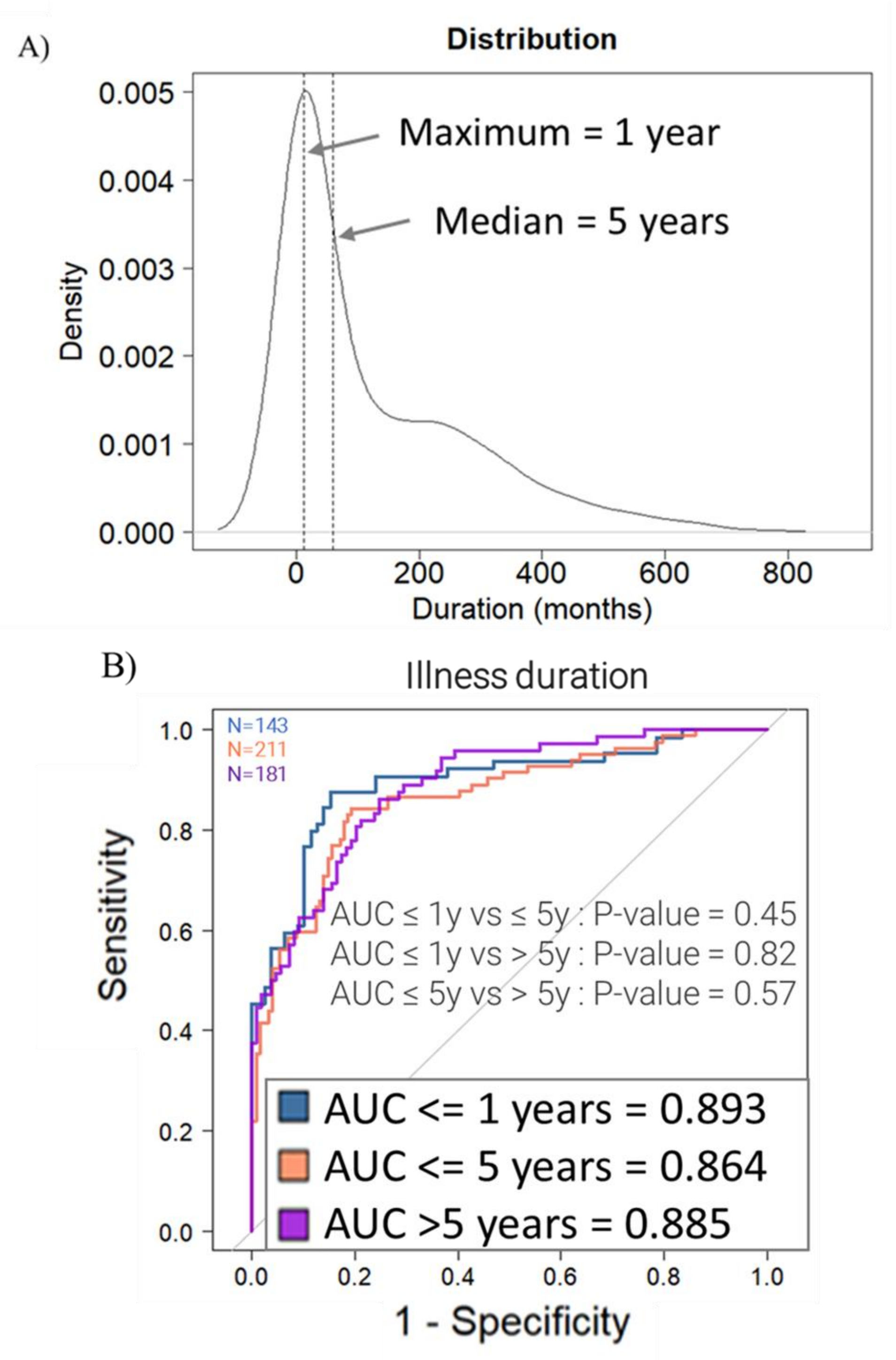

**Supplementary Figure15. Stratified sensitivity analysis according to number of depression episode.** A) Density plot showing the distribution of the number of depressive episodes in the analytical set (n=393), with a median of 4 episodes and a peak (mode) at 3 episodes. (B) Receiver operating characteristic (ROC) curves comparing the diagnostic performance of EDIT-B for patients with ≤ 3 depressive episodes (blue; AUC = 0.87) versus those with >3 episodes (coral; AUC = 0.849). (C) ROC curves comparing performance for patients with ≤4 depressive episodes (blue; AUC = 0.881) versus those with >4 episodes (coral; AUC = 0.836). In both analyses, no significant difference in area under the curve (AUC) was observed (p = 0.71 and p = 0.35, respectively), indicating that the discriminative power of the EDIT-B test is independent of the number of prior depressive episodes.

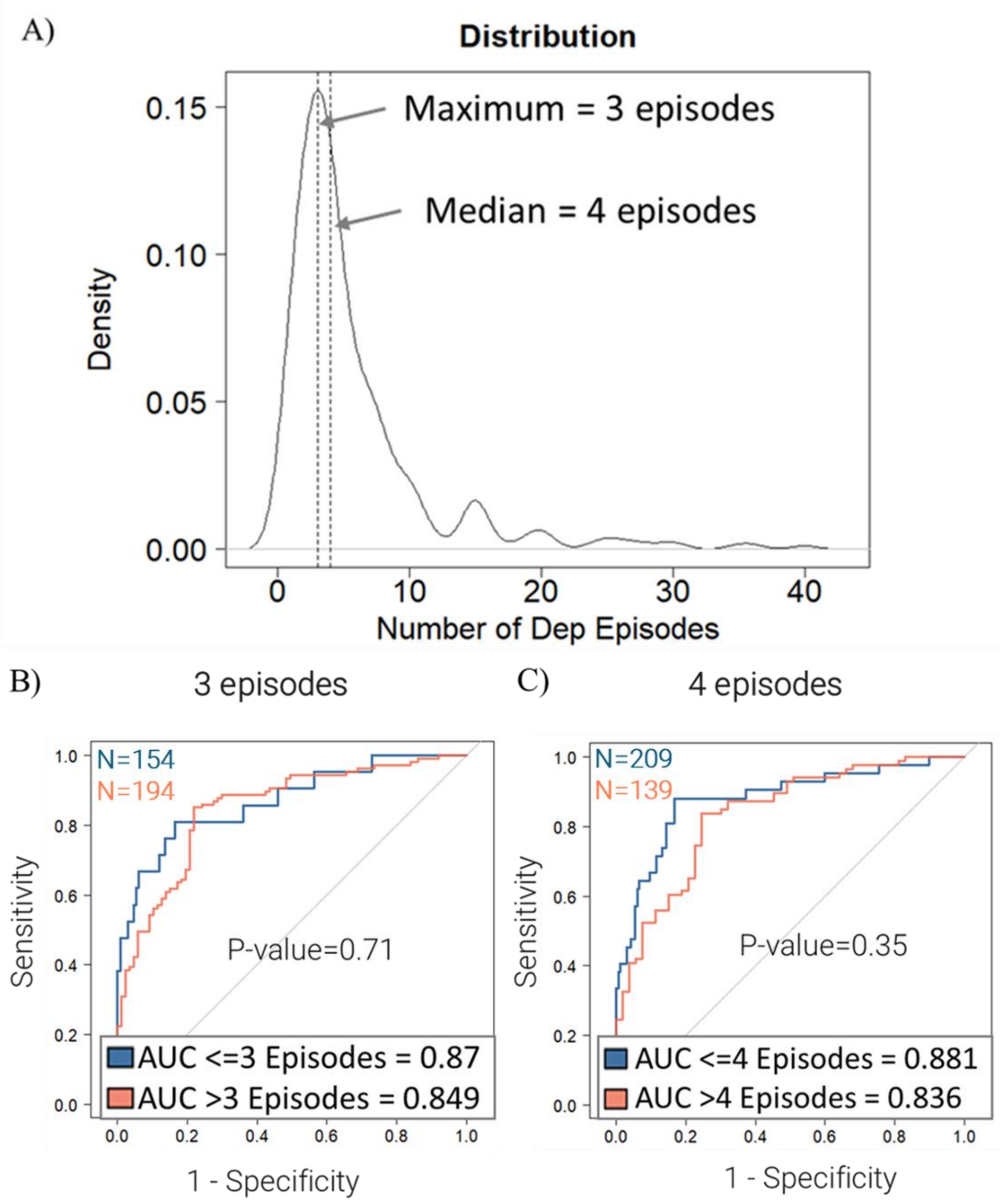

**Supplementary Figure16. EDIT-B True Positive (TP) and False Negative (FN) rates according to the number of manic, hypomanic, and mixed episodes.** The bar charts illustrate the proportion of True Positives (TP, blue) and False Negatives (FN, yellow) among patients with bipolar disorder, categorized by the number of prior episodes. **(A)** Performance relative to the number of manic episodes. **(B)** Performance relative to the number of hypomanic episodes. **(C)** Performance relative to the number of mixed episodes. Notably, the True Positive rate remains high across all strata, and the False Negative rate decreases toward zero as the number of episodes increases, demonstrating that the EDIT-B test's accuracy is maintained regardless of the frequency of manic/hypomanic/mixed episodes.

A)

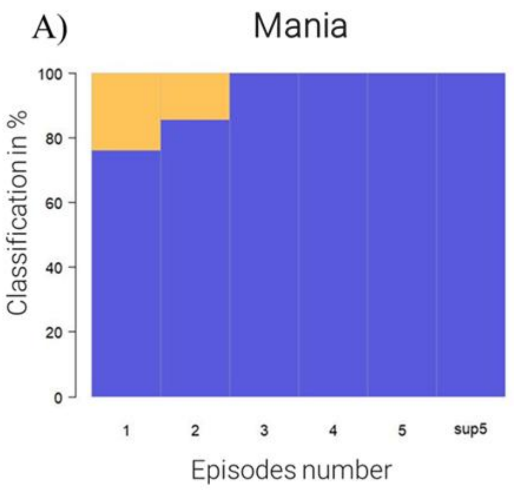


| N | 1 | 2 | 3 | 4 | 5 | >5 |
|---|---|---|---|---|---|---|
| TP (BD) | 16 | 12 | 4 | 4 | 2 | 7 |
| FN (MDD) | 5 | 2 | 0 | 0 | 0 | 0 |

B)

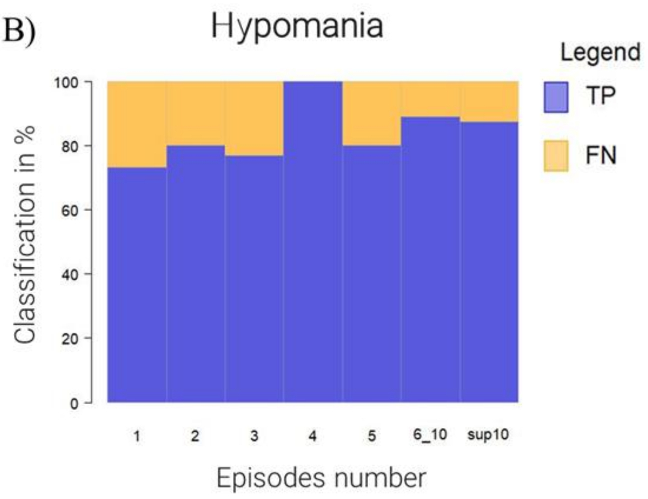


| N | 1 | 2 | 3 | 4 | 5 | [6-10] | >10 |
|---|---|---|---|---|---|---|---|
| TP (BD) | 11 | 16 | 10 | 9 | 8 | 16 | 7 |
| FN (MDD) | 4 | 1 | 3 | 0 | 2 | 2 | 1 |

C)

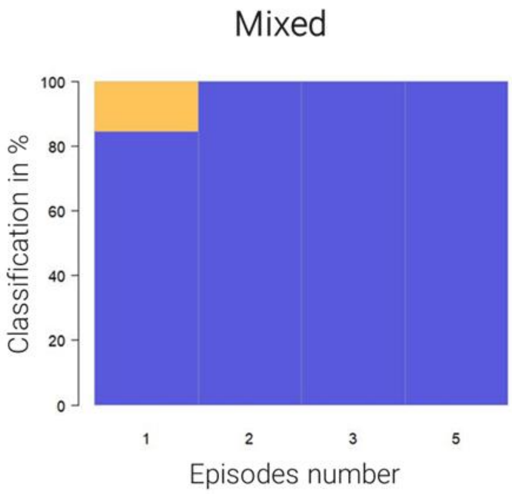


| N | 1 | 2 | 3 | 4 | 5 |
|---|---|---|---|---|---|
| TP (BD) | 11 | 5 | 1 | 0 | 3 |
| FN (MDD) | 2 | 0 | 0 | 0 | 0 |

**Supplementary Figure17. Stratified sensitivity analysis according to blood collection day-times.** (A) Distribution of blood collection times for MDD (orange), BD (red), and all patients (black). (B) ROC curves comparing EDIT-B performance for samples collected before 11:00 AM (AUC = 0.91) and after 11:00 AM (AUC = 0.871). No significant difference in diagnostic accuracy was observed between the two time periods (p = 0.31), demonstrating the stability of the EDIT-B test regardless of the sampling hour.

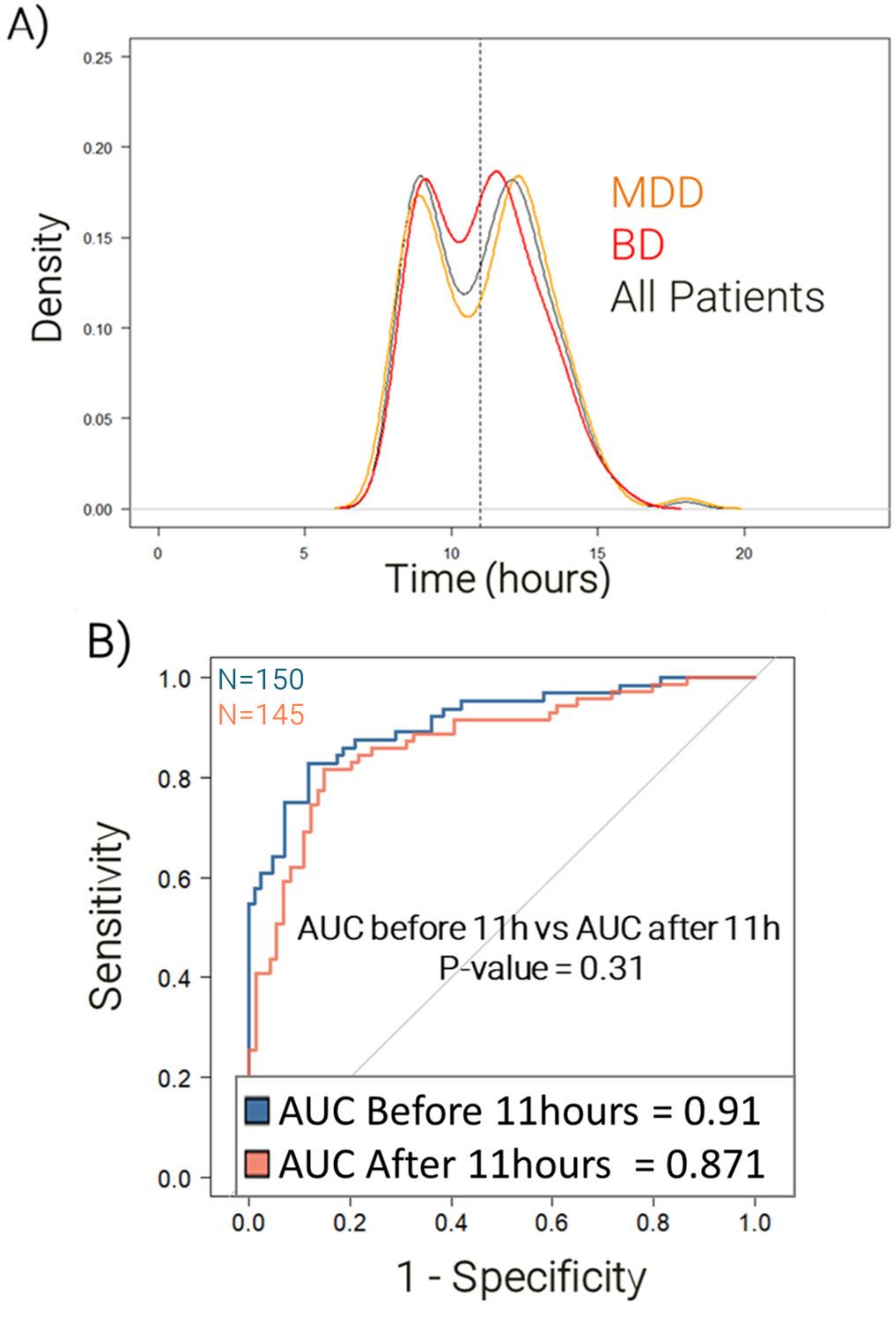

**Supplementary Figure18. Correlation analysis between RNA editing biomarkers and clinical covariates.** Heatmap representing the Pearson correlation coefficients between the 22 RNA editing biomarkers and a comprehensive set of patient covariates, including psychiatric medications, demographic data (age, sex, BMI, race), clinical scores (MADRS, YMRS), and disease characteristics (illness duration, age of onset, episode frequency, and psychiatric comorbidities). The predominantly neutral or light color scale indicates weak correlations across all variables, confirming that the EDIT-B molecular signatures are not significantly influenced by these potential confounding factors.

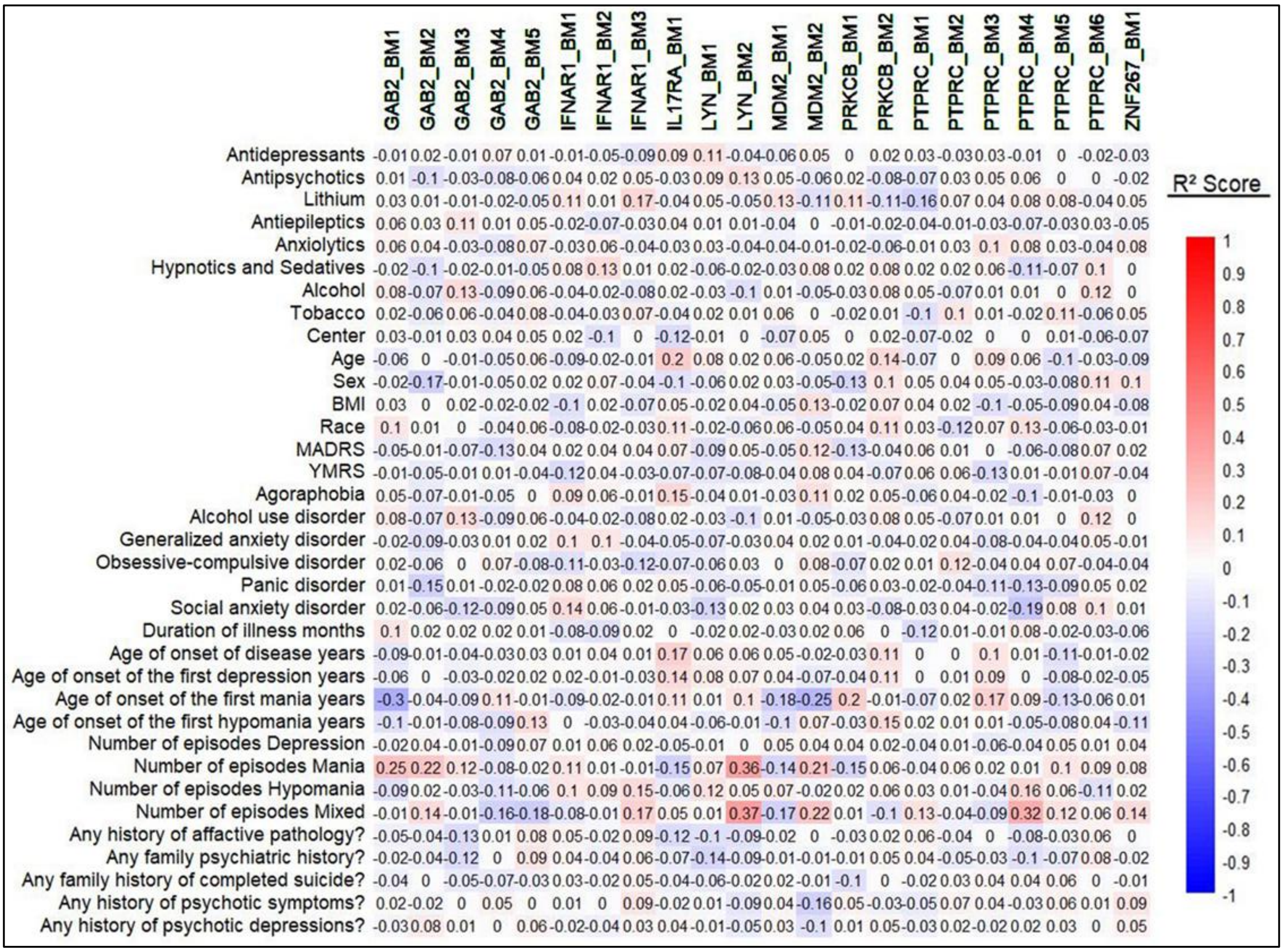

**Supplementary Figure19. Decision Curve Analysis for the diagnostic performance of the EDIT-B model.** The blue line represents the net benefit of the EDIT-B model in differentiating bipolar disorder from unipolar depression (shaded area indicates the 95% confidence interval), compared to the strategies of classifying all patients as bipolar (grey dashed line) or none as bipolar (black horizontal dotted line). The model consistently outperformed the assumption that "all" or "none" patients have bipolar disorder. The net benefit of the EDIT-B model remained significantly above the 'all bipolar' curve across most clinically relevant thresholds (from ≈ 0.2 to 0.9). The fact that the 95% confidence intervals remained entirely above the 'none bipolar' baseline suggests that the EDIT-B model can effectively reduce diagnostic errors (specifically the over-diagnosis of bipolar disorder) while maintaining high sensitivity for the detection of bipolarity.

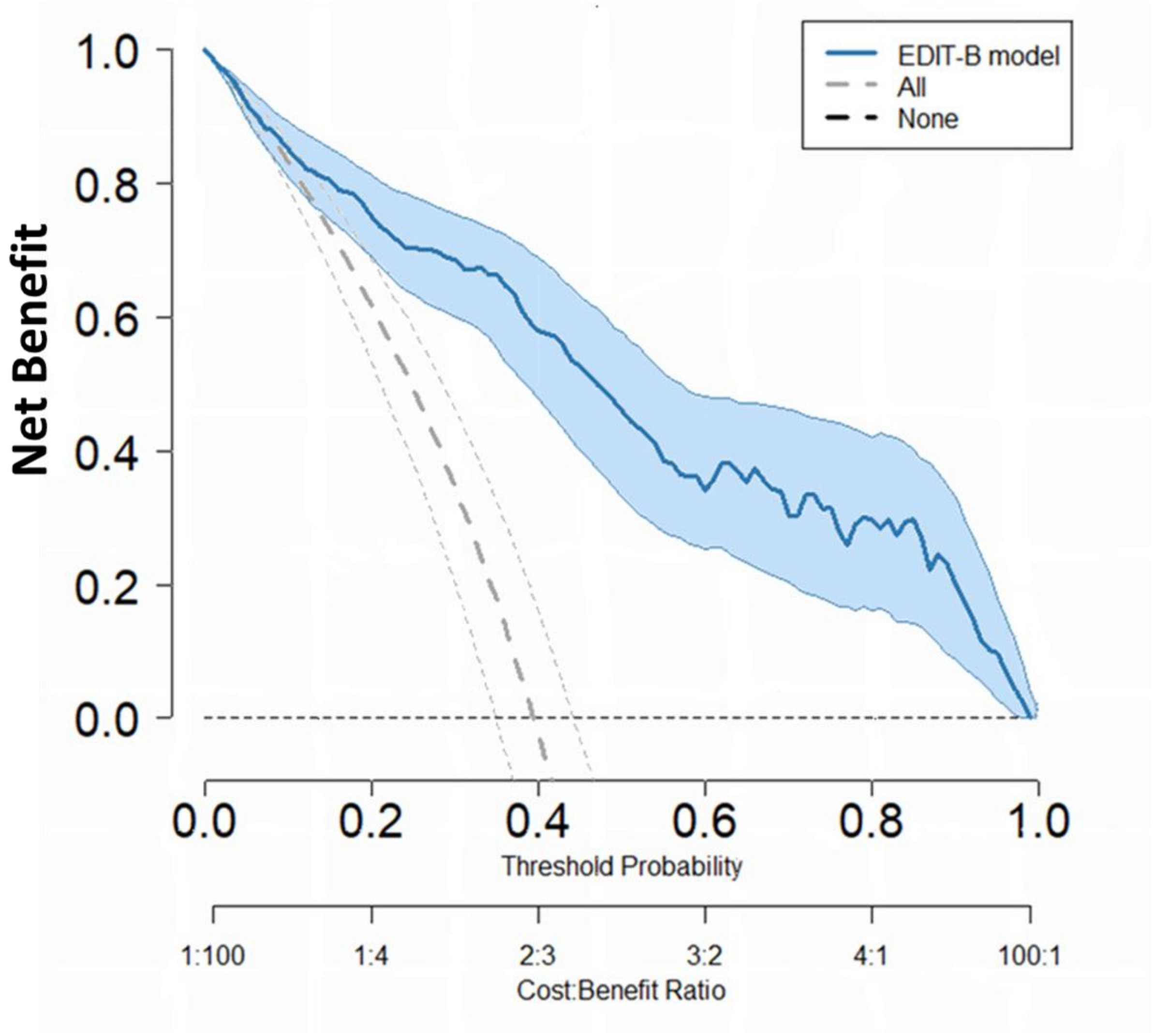
